# Cognitive subtypes in youth at clinical high risk for psychosis

**DOI:** 10.1101/2024.08.07.24311240

**Authors:** Walid Yassin, James Green, Matcheri Keshavan, Elisabetta C. del Re, Jean Addington, Carrie E Bearden, Kristin S Cadenhead, Tyrone D Cannon, Barbara A Cornblatt, Daniel H Mathalon, Diana O Perkins, Elaine F Walker, Scott W Woods, William S. Stone

## Abstract

**Introduction:** Schizophrenia is a mental health condition that severely impacts well-being. Cognitive impairment is among its core features, often presenting well before the onset of overt psychosis, underscoring a critical need to study it in the psychosis proneness (clinical high risk; CHR) stage, to maximize the benefits of interventions and to improve clinical outcomes. However, given the heterogeneity of cognitive impairment in this population, a one-size-fits-all approach to therapeutic interventions would likely be insufficient. Thus, identifying cognitive subtypes in this population is crucial for tailored and successful therapeutic interventions. Here we identify, validate, and characterize cognitive subtypes in large CHR samples and delineate their baseline and longitudinal cognitive and functional trajectories.

**Methods:** Using machine learning, we performed cluster analysis on cognitive measures in a large sample of CHR youth (n = 764), and demographically comparable controls (HC; n = 280) from the North American Prodrome Longitudinal Study (NAPLS) 2, and independently validated our findings with an equally large sample (NAPLS 3; n = 628 CHR, 84 HC). By utilizing several statistical approaches, we compared the clusters on cognition and functioning at baseline, and over 24 months of followup. We further delineate the conversion status within those clusters.

**Results:** Two main cognitive clusters were identified, “impaired” and “intact” across all cognitive domains in CHR compared to HC. Baseline differences between the cognitively intact cluster and HC were found in the verbal abilities and attention and working memory domains. Longitudinally, those in the cognitively impaired cluster group demonstrated an overall floor effect and did not deteriorate further over time. However, a “catch up” trajectory was observed in the attention and working memory domain. This group had higher instances of conversion overall, with these converters having significantly more non-affective psychotic disorder diagnosis versus bipolar disorder, than those with intact cognition. In the cognitively intact group, we observed differences in trajectory based on conversion status, where those who start with intact cognition and later convert demonstrate a sharp decline in attention and functioning. Functioning was significantly better in the cognitively intact than in the impaired group at baseline. Most of the cognitive trajectories demonstrate a positive relationship with functional ones.

**Conclusion:** Our findings provide evidence for intact and impaired cognitive subtypes in youth at CHR, independent of conversion status. They further indicate that attention and working memory are important to distinguish between the CHR with intact cognition and controls. The cognitively intact CHR group becomes less attentive after conversion, while the cognitively impaired one demonstrates a catch up trajectory on both attention and working memory. Overall, early evaluation, covering several cognitive domains, is crucial for identifying trajectories of improvement and deterioration for the purpose of tailoring intervention for improving outcomes in individuals at CHR for psychosis.

## Introduction

Schizophrenia is a debilitating mental health condition that affects overall well-being and is ranked among the top causes of disability-adjusted life years (1–4). The disability weight for an acute psychotic state is estimated to be the highest of any psychiatric condition (5). Schizophrenia is characterized by positive, negative, and other symptoms in addition to impaired cognition (6–8). Cognitive impairment in this population includes challenges with attention, verbal and working memory, executive functioning, and processing speed, among others (9–11). This has been demonstrated to significantly impact daily functioning (9–12). Given the chronicity of schizophrenia, focus is now shifting towards those in the early phase of the condition, clinical high risk for psychosis (CHR), during teenage years and early adulthood, when intervention is key (13,14). Individuals at CHR also experience varying levels of disruptions in perceptual and cognitive processes, as well as functioning, leading to worse overall outcomes (15–17).

Cognitive variability also characterizes individuals with first-episode psychosis (FEP) (18), and other psychotic disorders (19–21). Individuals who develop bipolar disorder, for example, typically show better overall cognition than those who develop schizophrenia (22–24). Several studies report cognitive deficits in the CHR population similar to those observed in other psychotic conditions, albeit milder, and these deficits are also quite heterogeneous (22,25–27). This heterogeneity hinders efforts for tailored therapeutic interventions (19). It is hoped that individuals at CHR will benefit from early intervention that would ameliorate symptoms and even prevent conversion to frank psychosis (28,29). This presents a valuable public health target and an opportunity to substantially improve the wellbeing of those impacted.

The challenges addressed above, among others, prompted a search for biomarkers to enhance the accuracy of diagnosis, prognosis, and treatment using machine learning (ML) and advanced statistical approaches (19,30–33). Cognition is among these biomarkers (17,22,27,34–37).

Given the functional importance of cognitive deficits in this population and the heterogeneity of their presentation, the identification of cognitive clusters may reveal subtypes of individuals who are most likely to benefit from tailored therapeutic interventions to achieve better outcomes (27,29,38,39).

Several studies attempted to produce cognitive subtypes in the CHR population, however, these studies either included other populations, such as genetic high-risk, FEP, recent onset psychosis or depression, controls (22,25), or other input variables such as clinical measures in addition to cognition (18,19,35). Moreover, the samples from the majority of these studies are relatively small with no external validation sample. The cognitive baseline and longitudinal trajectories in the CHR groups within different conversion statuses as well as their associated clinical and functional outcomes have not yet been reported.

The goal of this study is to identify, validate, and characterize cognitive subtypes in a large CHR population and to evaluate how these subtypes differ in clinical and functional outcomes. In addition, we explore these subtypes within converters and non-converters. To do this, we utilized the North American Prodrome Longitudinal Study (NAPLS) 2 neurocognitive data, clustered them using machine learning, and validated our findings independently using NAPLS 3. We then compared the clusters on different cognitive and functional measures.

## Methods

### Participants

The participants’ data in this study were obtained from NAPLS 2 (CHR (n = 764) HC (n = 280)) and 3 (CHR (n = 628) HC (n = 84)) consortia (13,14,40). Participants were help-seeking individuals who were referred from multiple community-based sources. Some were also self-referred. All CHR participants met the Criteria of Prodromal Syndrome (COPS) which is an assessment based on the Structured Interview for Psychosis-Risk Syndromes (SIPS) (41). After a thorough assessment which included administering the Structured Clinical Interview for DSM (SCID) IV (42) for NAPLS 2 and SCID 5 for NAPLS 3 (43), and the SIPS, vignettes were established for each CHR participant to obtain a consensus diagnosis. The attenuated psychotic symptoms rated on the Scale of Psychosis-Risk Symptoms (SOPS) are comprehensively described and include both recent and long-standing symptoms. The cross-site reliability in the SOPS rating was conducted in the same manner, with NAPLS 2 having an intraclass correlation range for the total SOPS of 0.82 to 0.93 and NAPLS 3 of 0.83 to 0.91, while the attenuated psychotic symptom score from the SOPS ranged from 0.92 to 0.96 and 0.83 to 0.94 for NAPLS 2 and 3, respectively. The differences across the individual sites were minimal and had excellent interclass correlations range. The Institutional Review Board approval was abstained from all sites and all the participants signed an informed consent.

### Inclusion and exclusion criteria

Participants in NAPLS 2 and 3 were between 12 and 35 years old and met the diagnostic criteria for the psychosis risk syndrome as part of the COPS criteria (41). Participants were excluded for i) criteria for current or lifetime Axis I psychotic disorder (Including affective psychosis), ii) IQ < 70, iii) history of disorders of the central nervous system, iv) diagnostic psychosis risk symptoms better explained by another Axis I disorder. Unless the disorder did not account for the individual’s prodromal symptoms, other non-psychotic DSM-IV and 5 disorders were not exclusionary, e.g., substance use disorder (13,14,40). The use of antipsychotics was not exclusionary if there was clear evidence that psychosis risk, not psychotic symptoms, were present when the medication was started. The healthy controls (HC) did not meet the criteria for any psychosis risk syndrome, a current or past psychotic disorder, or a Cluster A personality disorder diagnosis. In addition, we excluded those who are currently using psychotropic medication, as well as those with first-degree relatives having a history of a psychotic disorder or other disorders involving psychotic symptoms. Detailed information about the recruitment, diagnostic measures, and inclusion and exclusion criteria can be found elsewhere (13,14,40).

### Assessments

#### Clinical

Clinical measures included in this study were the SOPS, Global Assessment of Functioning (GAF) (44), and the Global Functioning Social (GF:SS) and Role Scales (GF:RS) (45).

#### Neurocognition

Neurocognitive measures that are common between NAPLS 2 and 3 were used. For example, newer versions of tests used in NAPLS2 were used in NAPLS 3, such as the Wechsler Abbreviated Scale of Intelligence (WASI) in NAPLS2 and the WASI-II in NAPLS 3 (46). In both instances, IQ was estimated from two subtests that included Vocabulary, but NAPLS2 used Block Design as the second test and NAPLS3 used Matrix Reasoning. Thus, block design and matrix reasoning were not included, only Vocabulary was. Similarly, portions of Seidman’s Auditory Continuous Performance (A-CPT) that were administered in NAPLS2 on cassette tapes were repeated in NAPLS3 but in a digitized format and administered by computer.

Portions of the A-CPT that were used in both NAPLS2 and NAPLS3 were included in the study and comprised the vigilance (QA) and high working memory load/no interference (Q3A-MEM) conditions (47), as was The Wide Range Achievement Test, Fourth Edition (WRAT-4) (48), and several MATRICS Consensus Cognitive Battery (MCCB) tests, including Brief Assessment of Cognition in Schizophrenia Symbol Coding (BACS SC) (49), Hopkins Verbal Learning Test-Revised (HVLT-R) (50), Letter-Number Span (LNS) (51).

#### Magnetic Resonance Imaging

The Magnetic Resonance Imaging scans were acquired using 3 Telsa Siemens (12-channel coil used in 5 sites) or a General Electric (8-channel coil used in 3 sites) scanners. The parameters of the sequences were optimized for the scanner manufacturer, version of the software, and coil configuration (http://adni.loni.ucla.edu/research/protocols/mri-protocols/) (52). Segmentation was performed using the Freesurfer software suite version 5.3. (53,54). Further details can be found elsewhere (55,56).

#### Conversion to psychosis

Conversion to psychosis was established by meeting the Presence of Psychotic Symptoms (POPS) criteria (57). At least one of the 5 SOPS positive symptoms reached a psychotic level of intensity, rated 6, for a frequency of more than one hour/day for 4 days/week during the past month, or symptoms seriously impacted functioning were needed to determine conversion.

### Machine learning

#### Preprocessing

NAPLS 2 and 3 were processed in the same way. Instances with more than 50% missingness of cognitive data were not included in the study. NAPLS 2 functioned as a discovery dataset and NAPLS 3 as a validation dataset. Only neurocognitive variables that were common to both consortia were included as input variables for clustering (See *Neurocognition* section).

Clustering was applied to CHR only (i.e., not healthy controls), since our aim was to identify cognitive subtypes within the CHR population. First, NAPLS 2 data were imputed using KKNImputer, part of scikit learn v1.4.2 (https://scikit-learn.org/), then the imputation of NAPLS 2 was applied to NAPLS 3 using the imputer fit/transofrm functions. Site correction was done using ComBat Family of Harmonization Methods (https://github.com/andy1764/ComBatFamily) in R Studios v4.3.2. The site effect removal was also applied to NAPLS 3 data using the function provided in ComBat. After, the age and sex effects were removed from both NAPLS 2 and 3 using the control data with Python v3.11.5 via scikit learn’s linear regression model. This was accomplished using regression to model the beta coefficients in the control group, to preserve illness effects, and to use them to residualize the cognitive variables in NAPLS 2 and 3 (13,14,40). Then, the data were standardized using the StandardScalar preprocessing function of scikit learn. Again, the standardization was done on NAPLS 2 and applied to NAPLS 3 using the fit-transform function. Lastly, principal component analysis (PCA) was used to reduce the multicollinearity of the data in NAPLS 2 using the decomposition package in scikit learn and applied to NAPLS 3.

#### Clustering algorithms

The clustering was done using the R package clValid v0.7 (58). The resulting principal components with 86.8% explained variance from the PCA entered the clustering. An in-house script was written to iterate over several algorithms (including k means, Partition Around Medoids (PAM), AGglomerative NESting (Agnes), and DIvisive ANAlysis Clustering (DIANA)), and available parameters (including metrics; euclidian; correlation; manhattan, methods; average; single; complete; ward, cluster number range [2:9], was based on the literature with some leeway for potentially higher number of clusters) and produce results that were compared based on the Silhouette score, to determine the best algorithm, with the optimal parameters, and the number of clusters. After identifying the algorithm and optimal parameters, as well as the number of clusters, the cluster membership was extracted from that algorithm, and used for further analysis.

#### Validation

NAPLS 2 clusters were validated in three ways using NAPLS 3 as indicated in Ullmann et al. (59). i) using external validators. We used total brain volume as an external validator (19) since a reduction in brain volume has been observed in those with impaired cognition (60,61). ii) Methods based validation. The PCs from NAPLS 3 were run using the same algorithm, parameters, and cluster number that were deemed best for the NAPLS 2 data and selected to produce the NAPLS 3 clusters. Then, the two were compared based on the Silhouette score. A similar silhouette score is indicative of good validation (59). iii) Results based validation. The centroids of the clusters in NAPLS 2 were calculated and the instances in NAPLS 3 were mapped on those clusters, i.e., the proximity of each instance in NAPLS 3 to the centroid produced from the clusters of NAPLS 2 would determine their cluster membership (59). After, a machine learning classification algorithm (random forest classifier using sklearn) was run using the cluster membership as labels and the PCs of the instances as input variables from NAPLS 2 for training, and then the trained algorithm from NAPLS 2 was used for classifying NAPLS 3 data. The results were reported including precision, recall, and f1-score. In addition, a confusion matrix is also provided.

#### Clusters vs converter status

After the clusters were identified and described, we were interested in delineating the characteristics of the converters and non-converters belonging to each of the cognitive clusters.

### Statistical Analysis

Descriptive statistics, including chi-square and t-tests, were implemented to assess the differences across demographic variables. Then, for NAPLS2 (discovery sample), demographics were described for the clusters produced. The clusters within the converter and non-converter groups are also presented. To describe baseline differences in cognitive, clinical, and functional outcomes between the clusters and controls, Kruskal-Wallis test with post-hoc pairwise comparisons was conducted. Dunn’s test was used for post-hoc pairwise comparisons where needed. Longitudinal analyses were conducted using a linear mixed effects model in R, where the HC are used as reference. False Discovery Rate was used to correct for multiple comparisons. The comparisons of the clusters between the cognitive variables at baseline were the only analyses that were not corrected for multiple comparisons as they were produced synthetically and were presented for descriptive purposes. Kaplan-Meyer survival curve was used to identify differences in conversion between the clusters. This was conducted with the entire sample, however, another analysis was run with participants who remained in the study < 2.5 years to reduce outlier effects (62). The number of converters vs non -converters in the clusters was evaluated using the Chi square test. We assessed the longitudinal association between cognition and SOPS to further evaluate their longitudinal relationship post hoc, using a Generalized Estimating Equation (GEE). We also examine the relationship between cognition and functioning while accounting for the repeated measures within individual overtime, we used a GEE on the entire sample in R (63,64). Functioning was specified as outcome variables and cognition as predictors, and vice versa, both were scaled. The “waves” were the time points, and the participant ID was used to identify repeated measures for each participant. The correlation structures were chosen based on the quasi information criterion (QIC) (65). The “independence” correlation structure was not entered into the QIC model evaluation, as it is not suitable for this analysis. The percentage of psychotic vs bipolar diagnoses were calculated by using the total of the converters in each cluster as the total number of converters (e.g., of all the converters in C1, how many have a psychotic versus bipolar diagnosis?).

## Results

### Demographic data

The demographic data of NAPLS 2 and NAPLS 3 are presented in table S1. The demographic data of NAPLS 2 and 3 did not differ within each study between the controls and CHR for sex. Control participants in NAPLS 2 were about a year older (x̄ : 19.73 vs. 18.5) than CHR (p < 0.001). In both NAPLS2 and NAPLS3, CHR participants were more likely to have lower levels of education than the controls (p < 0.001). The resulting clusters did not differ in age and sex, but differ in income (p < 0.001), and education (p < 0.001). More details regarding other demographic factors can be found in table 1.

**Table 1:** Demographic comparisons for cluster membership.

|  | Mean (SD) or N (%) |  |  | p-value |
| --- | --- | --- | --- | --- |
|  | C1 (N=278) | C2 (N=322) | Statistic |  |
| Age | 18.53 (4.91) | 18.53 (3.67) | $t = 0.01$ | 0.988 |
| Sex, male: female | 148 (53.2%) | 194 (60.2%) | $\chi^2 = 2.71$ | 0.09 |
| Income | | | $\chi^2 = 37.28$ | < 0.001 |
| Less than 10,000 | 33 (12.0%) | 24 (7.5%) |  |  |
| 10,000 – 19,999 | 33 (12.0%) | 19 (5.9%) |  |  |
| 20,000 – 39,999 | 37 (13.5%) | 33 (10.3%) |  |  |
| 40,000 – 59,999 | 33 (12.0%) | 27 (8.4%) |  |  |
| 60,000 – 99,999 | 35 (12.7%) | 54 (16.8%) |  |  |
| 100,000 + | 32 (11.6%) | 94 (29.3%) |  |  |
| Don't know or refused | 72 (26.2%) | 70 (21.8%) |  |  |
| Education | | | $\chi^2 = 20.73$ | < 0.001 |
| No High School | 172 (62.1%) | 153 (47.7%) |  |  |
| High School | 86 (31.0%) | 128 (39.9%) |  |  |
| College | 7 (2.5%) | 14 (4.4%) |  |  |
| Technical school | 7 (2.5%) | 4 (1.2%) |  |  |
| University | 5 (1.8%) | 18 (5.6%) |  |  |
| Graduate school | 0 (0.0%) | 4 (1.2%) |  |  |

*C=cluster, SD = standard deviation, N=sample size*

### Machine learning

#### Clustering

The clustering analysis using NAPLS 2 (discovery) data demonstrated that DIANA was the best algorithm resulting in two clusters (Cluster 1, impaired (C1), Cluster 2, intact (C2)), with good separability indicated by the internal validation measures including the Silhouette score of 0.5045 (Table S2). Hierarchical clustering is a valuable overall clustering algorithm and has proven to be suitable for clustering cognition in CHR (22,25). The full details of the clustering results including parameters can be seen in the supplementary material (Table S2, S3). The clusters in 3D and dendrograms are also presented (Figures S1).

#### Validation

Total brain volume (mm^3^) was significantly different between the clusters (p=0.024), where C2 (Mean (M)=606,085, standard deviation (SD)=614,89) had a larger volume than C1 (M=592,433, SD=66,835), which is what we would expect based on evidence from the literature (19,60,61). NAPLS 3 clustering demonstrated a similar silhouette score (0.5408) to the NAPLS 2 (0.5045). The second validation method demonstrated an accuracy of 0.83 (Precision =0.88, recall=0.86, f1-score=.87) (See Table S3 for confusion matrix).

#### Cognition (Baseline)

As expected, there are significant differences in the cognitive domains that entered the clustering algorithm, where C1 is characterized by “impaired” cognition and C2 by “intact” cognition (WRAT (p <0.001), WASI Vocab (p <0.001), CPT QA (p < 0.001) and Q3A (p < 0.001), BACS (p < 0.001), HVLT-R (p < 0.001), LNS (p < 0.001)). Interestingly, only CPT Q3A (p <0.01) and WRAT (p < 0.05) show differences between C2 and controls (Table 2, Fig 1.)

**Figure 1:**
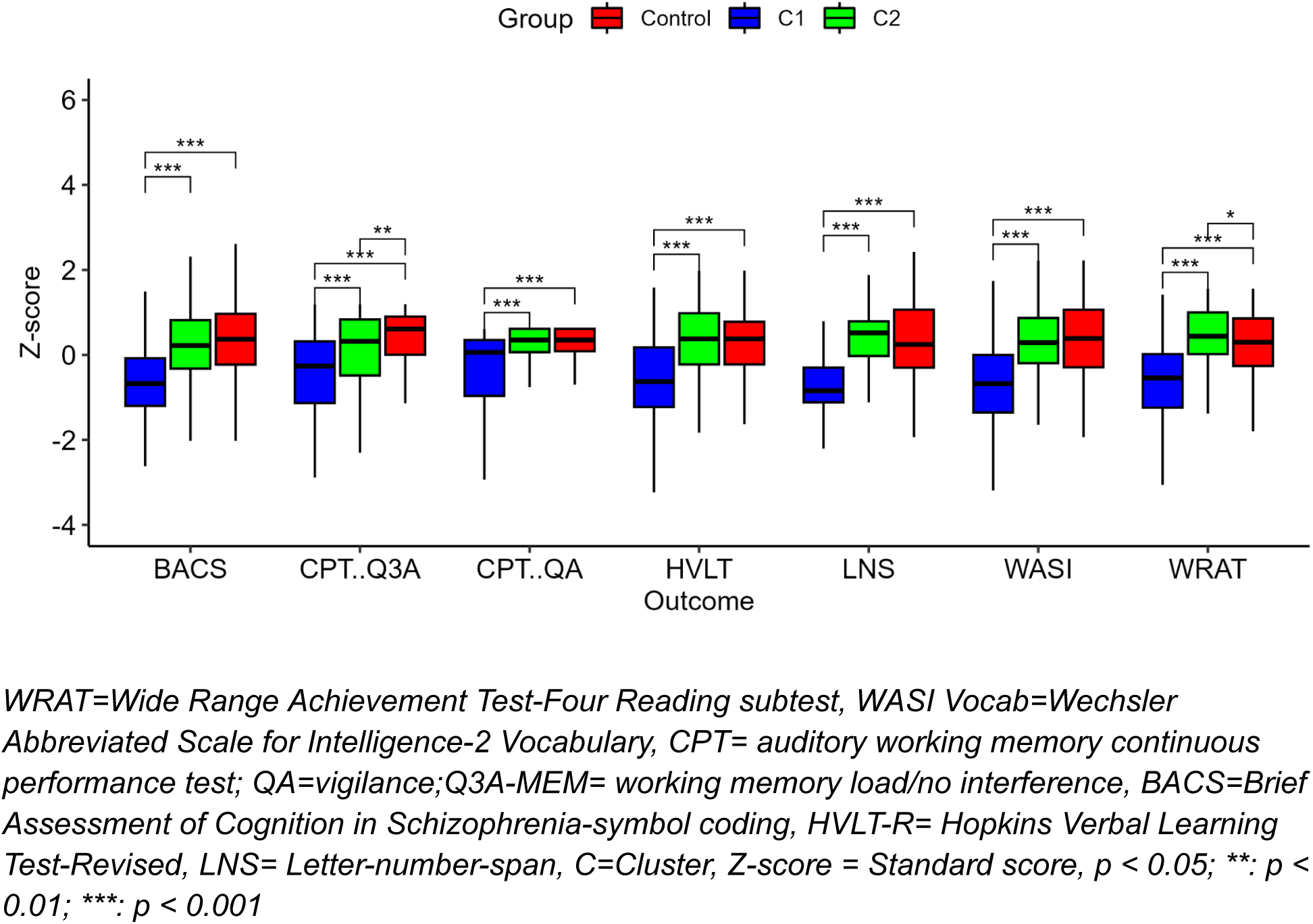
Showing the baseline cognitive differences between the clusters and controls

**Table 2:** Pairwise Comparisons between Cognitive Clusters, Controls, and Cognitive Outcomes.

a) Kruskal-Wallis Comparisons
| Outcome | C1 | C2 | Control | statistic | p-value |
| --- | --- | --- | --- | --- | --- |
| BACS | 50.12 (11.47) | 62.5 (11.29) | 64.27 (12.91) | $Z = 193.61$ | < 0.001 |
| HVLT | 22.96 (5.18) | 27.75 (4.03) | 27.44 (4.22) | $Z = 151.31$ | < 0.001 |
| LNS | 12.25 (3.2) | 16.56 (2.78) | 16.31 (3.37) | $Z = 252$ | < 0.001 |
| CPT: Q3A | 75.8 (17.14) | 85.02 (11.78) | 87.58 (10.82) | $Z = 90.48$ | < 0.001 |
| CPT: QA | 91.36 (10.91) | 97.06 (4.01) | 97.24 (4.93) | Z = 83.07 | < 0.001 |
| WASI | 48.84 (9.41) | 59.04 (8.49) | 59.82 (9.48) | Z = 192.52 | < 0.001 |
| WRAT | 53.92 (7.41) | 61.88 (4.97) | 60.39 (6.36) | Z = 204.9 | < 0.001 |

b) Dunn Pairwise Comparisons
| Outcome | Comparison (a, b) | Mean (SD) (a) | Mean (SD) (b) | Statistic | p-value |
| --- | --- | --- | --- | --- | --- |
| BACS | C1, C2 | 50.12 (11.47) | 62.5 (11.29) | Z = 11.5 | < 0.001 |
|  | C1, Control | 50.12 (11.47) | 64.27 (12.91) | Z = 12.61 | < 0.001 |
|  | C2, Control | 62.5 (11.29) | 64.27 (12.91) | Z = 1.68 | 0.093 |
| HVLT | C1, C2 | 22.96 (5.18) | 27.75 (4.03) | Z = 11.26 | < 0.001 |
|  | C1, Control | 22.96 (5.18) | 27.44 (4.22) | Z = 10.02 | < 0.001 |
|  | C2, Control | 27.75 (4.03) | 27.44 (4.22) | Z = -0.74 | 0.458 |
| LNS | C1, C2 | 12.25 (3.2) | 16.56 (2.78) | Z = 14.66 | < 0.001 |
|  | C1, Control | 12.25 (3.2) | 16.31 (3.37) | Z = 12.73 | < 0.001 |
|  | C2, Control | 16.56 (2.78) | 16.31 (3.37) | Z = -1.29 | 0.197 |
| CPT: Q3A | C1, C2 | 75.8 (17.14) | 85.02 (11.78) | Z = 6.76 | < 0.001 |
|  | C1, Control | 75.8 (17.14) | 87.58 (10.82) | Z = 9.24 | < 0.001 |
|  | C2, Control | 85.02 (11.78) | 87.58 (10.82) | Z = 2.88 | < 0.01 |
| CPT: QA | C1, C2 | 91.36 (10.91) | 97.06 (4.01) | Z = 7.64 | < 0.001 |
|  | C1, Control | 91.36 (10.91) | 97.24 (4.93) | Z = 8.23 | < 0.001 |
|  | C2, Control | 97.06 (4.01) | 97.24 (4.93) | Z = 0.93 | 0.3499 |
| WASI | C1, C2 | 48.84 (9.41) | 59.04 (8.49) | Z = 11.85 | < 0.001 |
|  | C1, Control | 48.84 (9.41) | 59.82 (9.48) | Z = 12.28 | < 0.001 |
|  | C2, Control | 59.04 (8.49) | 59.82 (9.48) | Z = 1.01 | 0.3105 |
| WRAT | C1, C2 | 53.92 (7.41) | 61.88 (4.97) | Z = 13.66 | < 0.001 |
|  | C1, Control | 53.92 (7.41) | 60.39 (6.36) | Z = 10.67 | < 0.001 |
|  | C2, Control | 61.88 (4.97) | 60.39 (6.36) | Z = -2.44 | < 0.05 |

*WRAT=Wide Range Achievement Test-Four Reading subtest, WASI Vocab=Wechsler Abbreviated Scale for Intelligence-2 Vocabulary, CPT= auditory working memory continuous performance test; QA=vigilance; Q3A-MEM= working memory load/no interference, BACS=Brief Assessment of Cognition in Schizophrenia-symbol coding, HVLT-R= Hopkins Verbal Learning Test-Revised, LNS= Letter-number-span, C=Cluster, SD=standard deviation.*

#### Cognition (Longitudinal)

C1 demonstrated a notable catch up trajectory in CPT QA (*b* = 2.17, *p* < 0.001), and Q3A (*b* = 3.03, *p* < 0.01), compared to controls. (Table 3, Fig 2). No other notable differences were observed.

**Figure 2:**
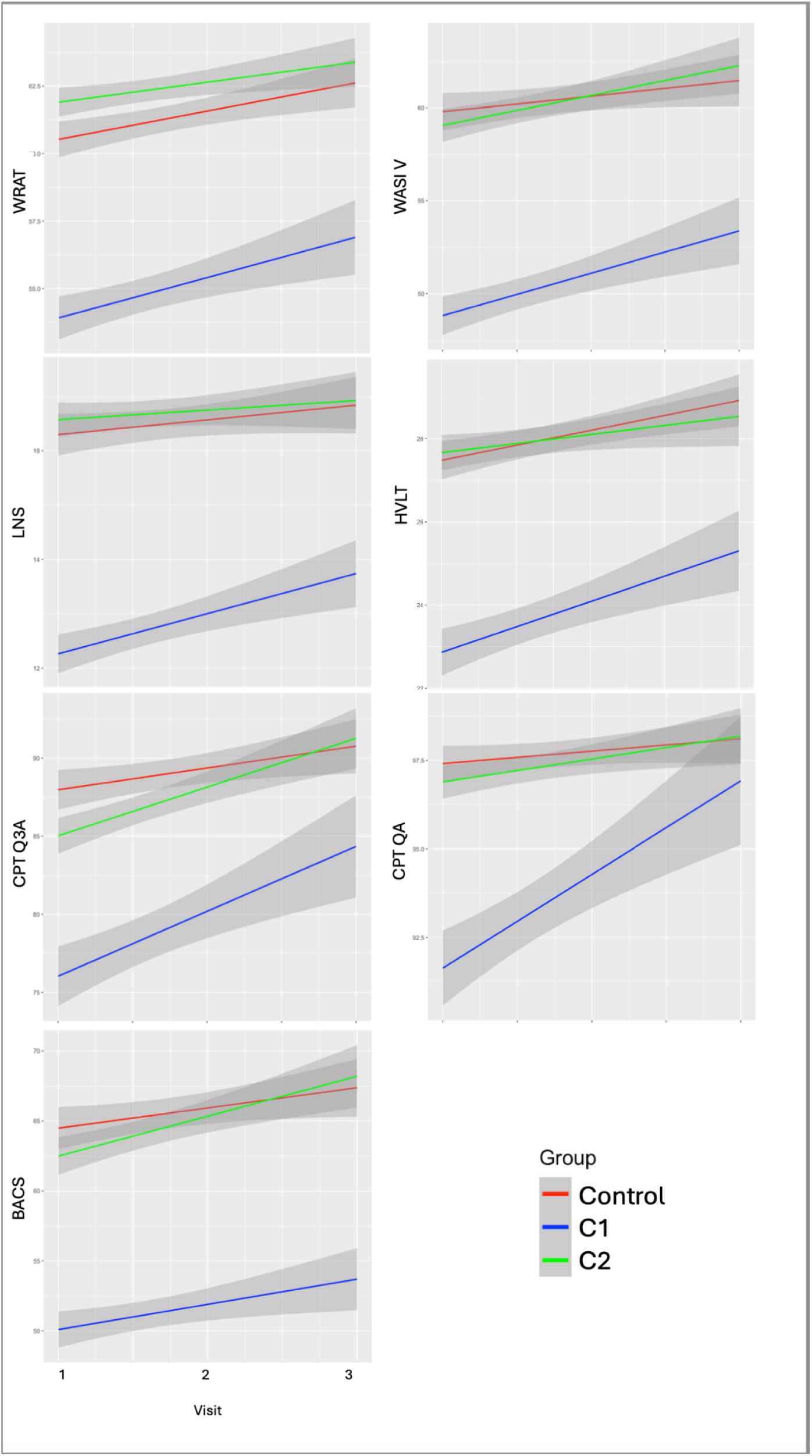
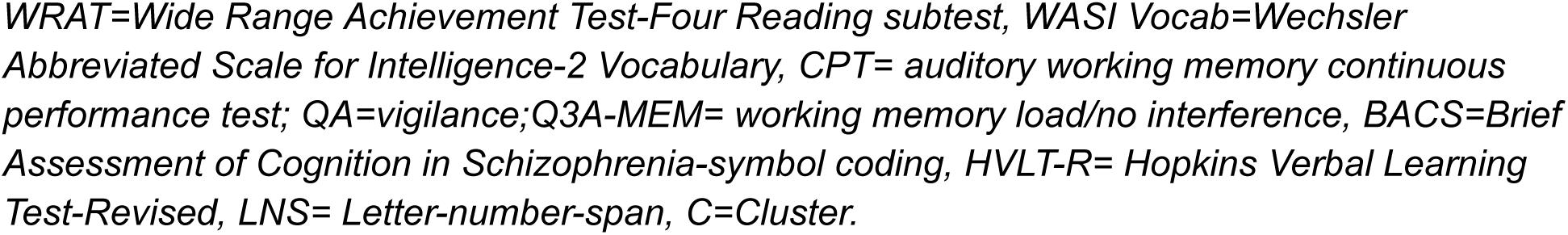
Longitudinal cognitive outcomes across clusters and controls.

**Table 3:** Cognitive Clusters and Longitudinal Cognition Outcomes.

|  |  | Estimate | Std.err | t | p-value |
| --- | --- | --- | --- | --- | --- |
| WRAT | (intercept) | 59.21 | 0.46 | 128.77 | < 0.001 |
|  | C1 | -6.91 | 0.66 | -10.54 | < 0.001 |
|  | C2 | 1.67 | 0.63 | 2.64 | < 0.01 |
|  | Visit | 1.23 | 0.15 | 8.32 | < 0.001 |
|  | C1*Visit | 0.41 | 0.23 | 1.75 | 0.09659 |
|  | C2*Visit | -0.30 | 0.22 | -1.35 | 0.177 |
| WASI: V | (intercept) | 58.50 | 0.69 | 85.02 | < 0.001 |
|  | C1 | -11.48 | 0.99 | -11.64 | < 0.001 |
|  | C2 | -0.94 | 0.95 | -0.99 | 0.3203 |
|  | Visit | 1.19 | 0.26 | 4.66 | < 0.001 |
|  | C1*Visit | 0.66 | 0.40 | 1.65 | 0.1501 |
|  | C2*Visit | 0.27 | 0.38 | 0.71 | 0.481 |
| CPT: QA | (intercept) | 97.02 | 0.64 | 151.13 | < 0.001 |
|  | C1 | -7.96 | 0.93 | -8.56 | < 0.001 |
|  | C2 | -0.72 | 0.89 | -0.82 | 0.4151 |
|  | Visit | 0.39 | 0.27 | 1.41 | 0.158 |
|  | C1*Visit | 2.17 | 0.42 | 5.16 | < 0.001 |
|  | C2*Visit | 0.23 | 0.40 | 0.58 | 0.561 |
| CPT: Q3A | (intercept) | 86.54 | 1.25 | 69.42 | < 0.001 |
|  | C1 | -15.06 | 1.82 | -8.26 | < 0.001 |
|  | C2 | -4.36 | 1.74 | -2.51 | < 0.05 |
|  | Visit | 1.38 | 0.60 | 2.32 | < 0.05 |
|  | C1*Visit | 3.04 | 0.93 | 3.26 | < 0.01 |
|  | C2*Visit | 1.50 | 0.88 | 1.71 | 0.087 |
| BACS | (intercept) | 62.51 | 0.92 | 68.19 | < 0.001 |
|  | C1 | -13.96 | 1.33 | -10.52 | < 0.001 |
|  | C2 | -1.68 | 1.27 | -1.32 | 0.1866 |
|  | Visit | 1.79 | 0.40 | 4.43 | < 0.001 |
|  | C1*Visit | -0.11 | 0.64 | -0.17 | 0.9863 |
|  | C2*Visit | 0.01 | 0.60 | 0.02 | 0.9865 |
| HVLT | (intercept) | 26.76 | 0.38 | 70.23 | < 0.001 |
|  | C1 | -4.75 | 0.55 | -8.62 | < 0.001 |
|  | C2 | 0.65 | 0.53 | 1.23 | 0.2178 |
|  | Visit | 0.69 | 0.17 | 4.04 | < 0.001 |
|  | C1*Visit | 0.22 | 0.27 | 0.81 | 0.4208 |
|  | C2*Visit | -0.41 | 0.25 | -1.59 | 0.1687 |
| LNS | (intercept) | 15.95 | 0.25 | 63.85 | < 0.001 |
|  | C1 | -4.33 | 0.36 | -11.99 | < 0.001 |
|  | C2 | 0.57 | 0.35 | 1.65 | 0.1002 |
|  | Visit | 0.34 | 0.11 | 3.08 | < 0.01 |
|  | C1*Visit | 0.32 | 0.18 | 1.81 | 0.0859 |
|  | C2*Visit | -0.30 | 0.17 | -1.81 | 0.085 |

*WRAT=Wide Range Achievement Test-Four Reading subtest, WASI Vocab=Wechsler Abbreviated Scale for Intelligence-2 Vocabulary, CPT= auditory working memory continuous performance test; QA=vigilance; Q3A-MEM= working memory load/no interference, BACS=Brief Assessment of Cognition in Schizophrenia-symbol coding, HVLTR= Hopkins Verbal Learning Test-Revised, LNS= Letter-number-span, C=Cluster, Std.err= Standard error.*

### Cognitive clusters vs. conversion status

#### Cognition (Baseline)

Pairwise comparisons demonstrated that controls and C2NC had higher scores than C1 independent of their conversion (p <0.001). C2C also had higher scores than C1 on all cognitive domains independent of conversion status (CPT Q3A: p<0.05, otherwise p<0.001).

The only differences between C2NC and controls were found in CPT Q3A and WRAT, where CPT Q3A was lower (p<0.05), and WRAT was higher (p<0.05) (Table 4, Fig 3).

**Figure 3:**
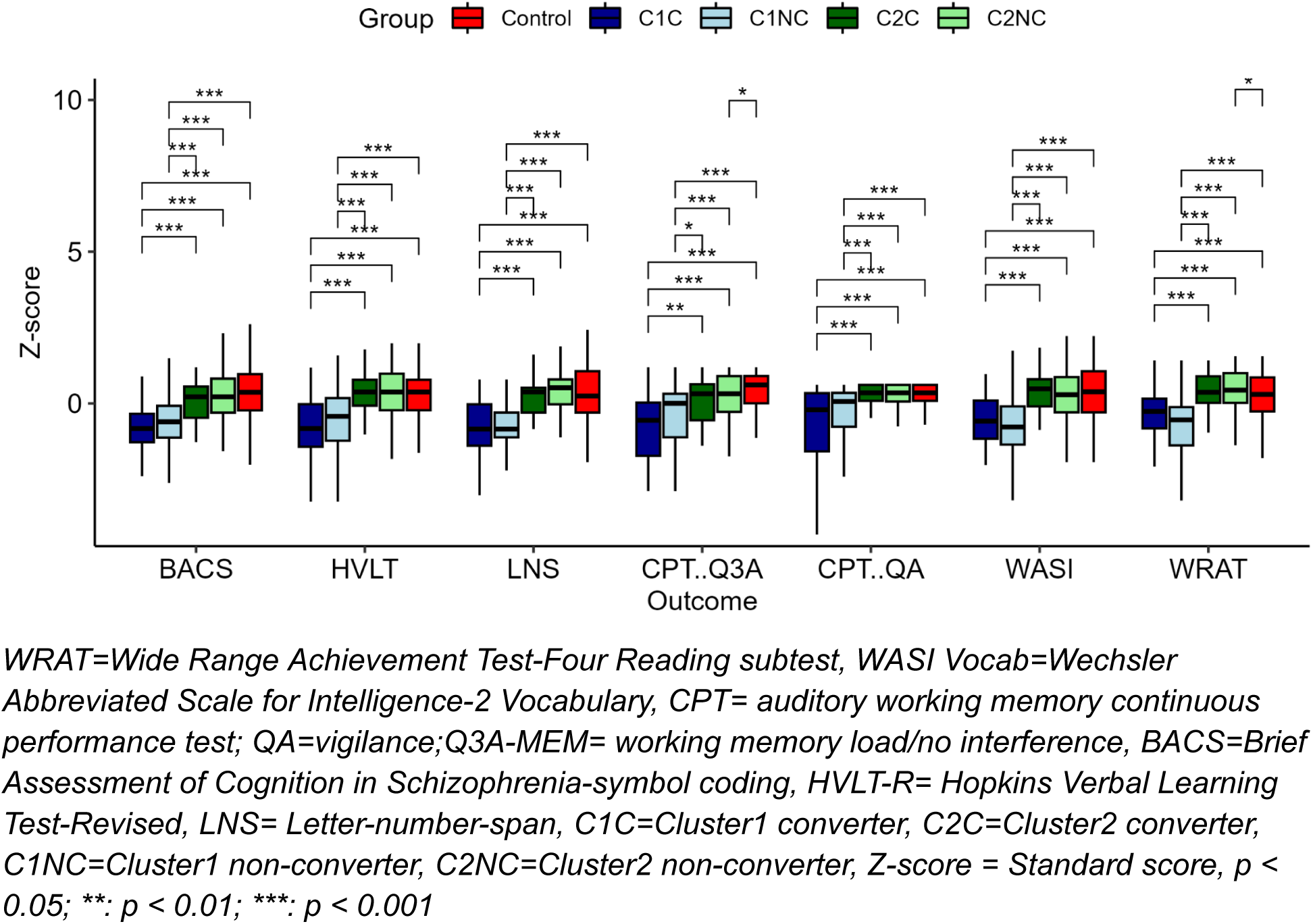
Pairwise comparisons between clusters, converters, and healthy controls.

**Table 4:**
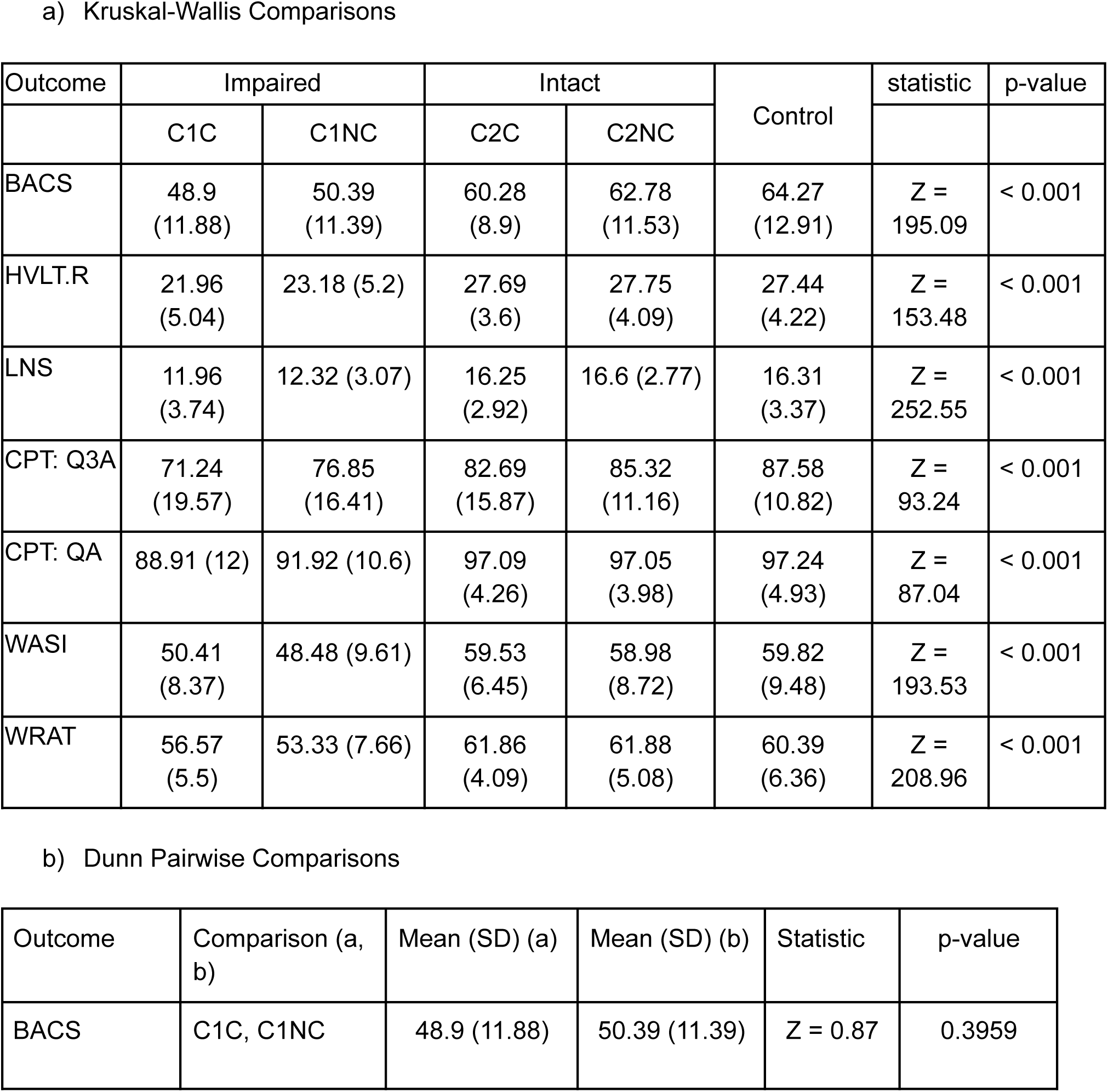
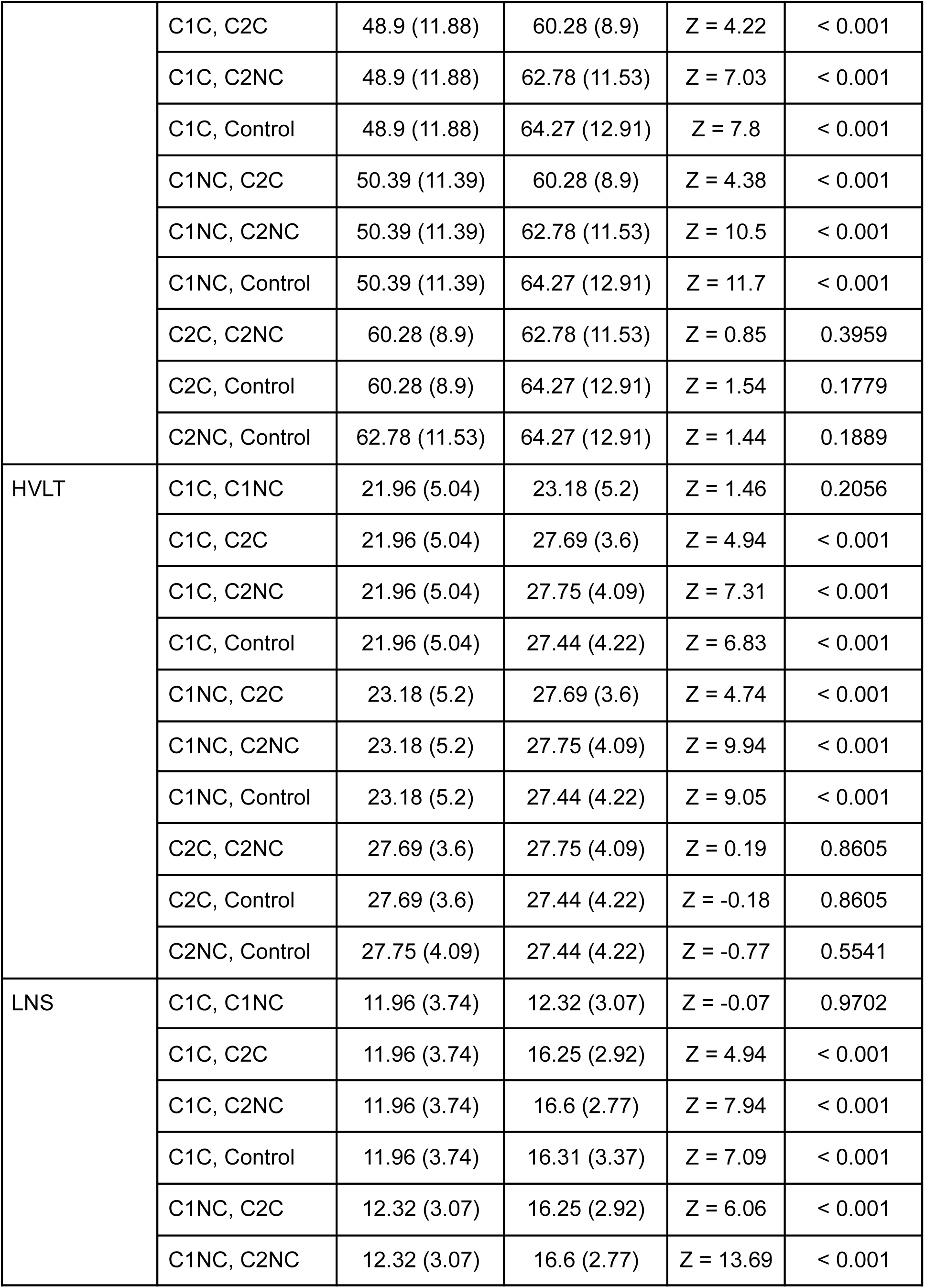
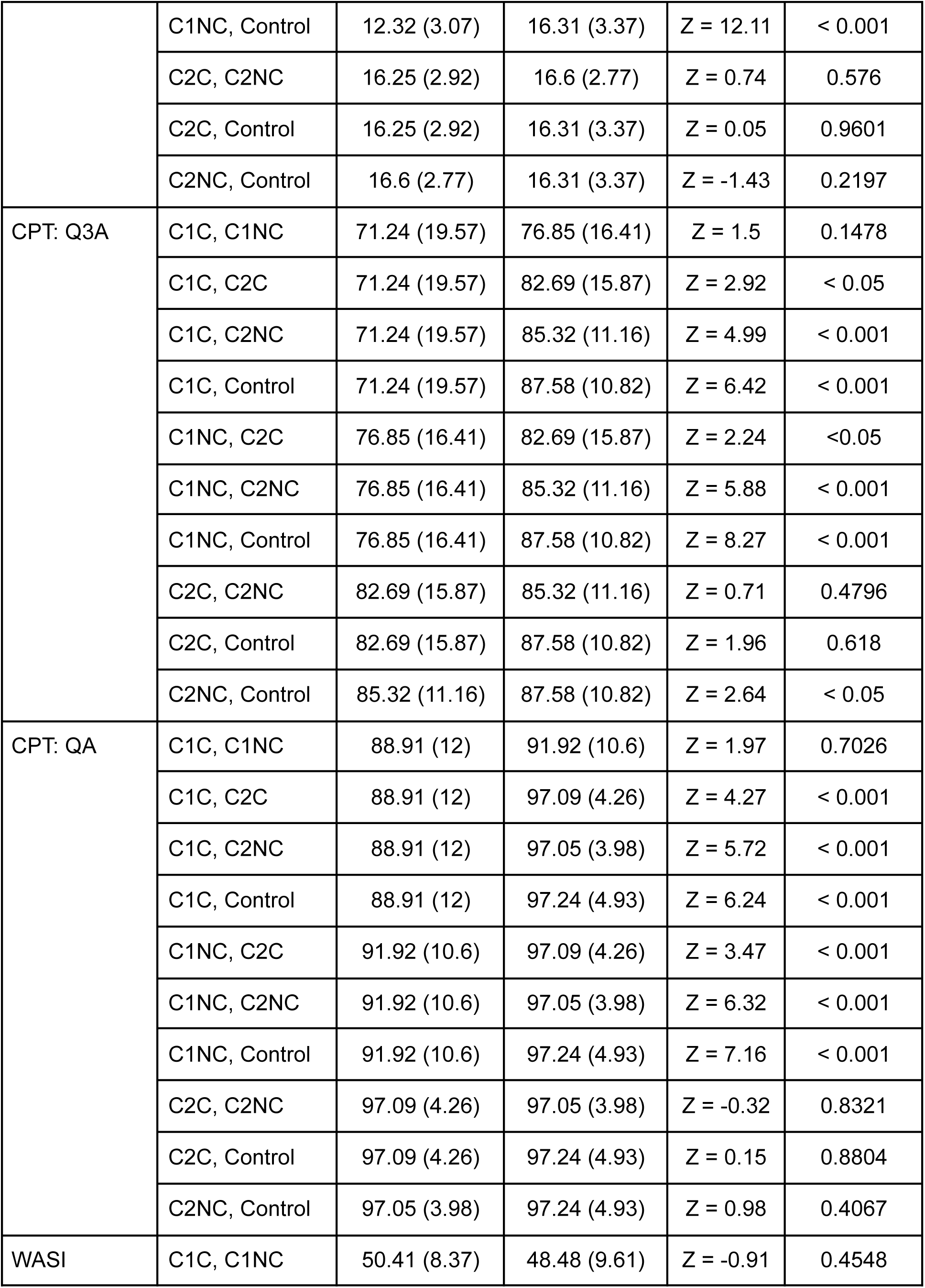
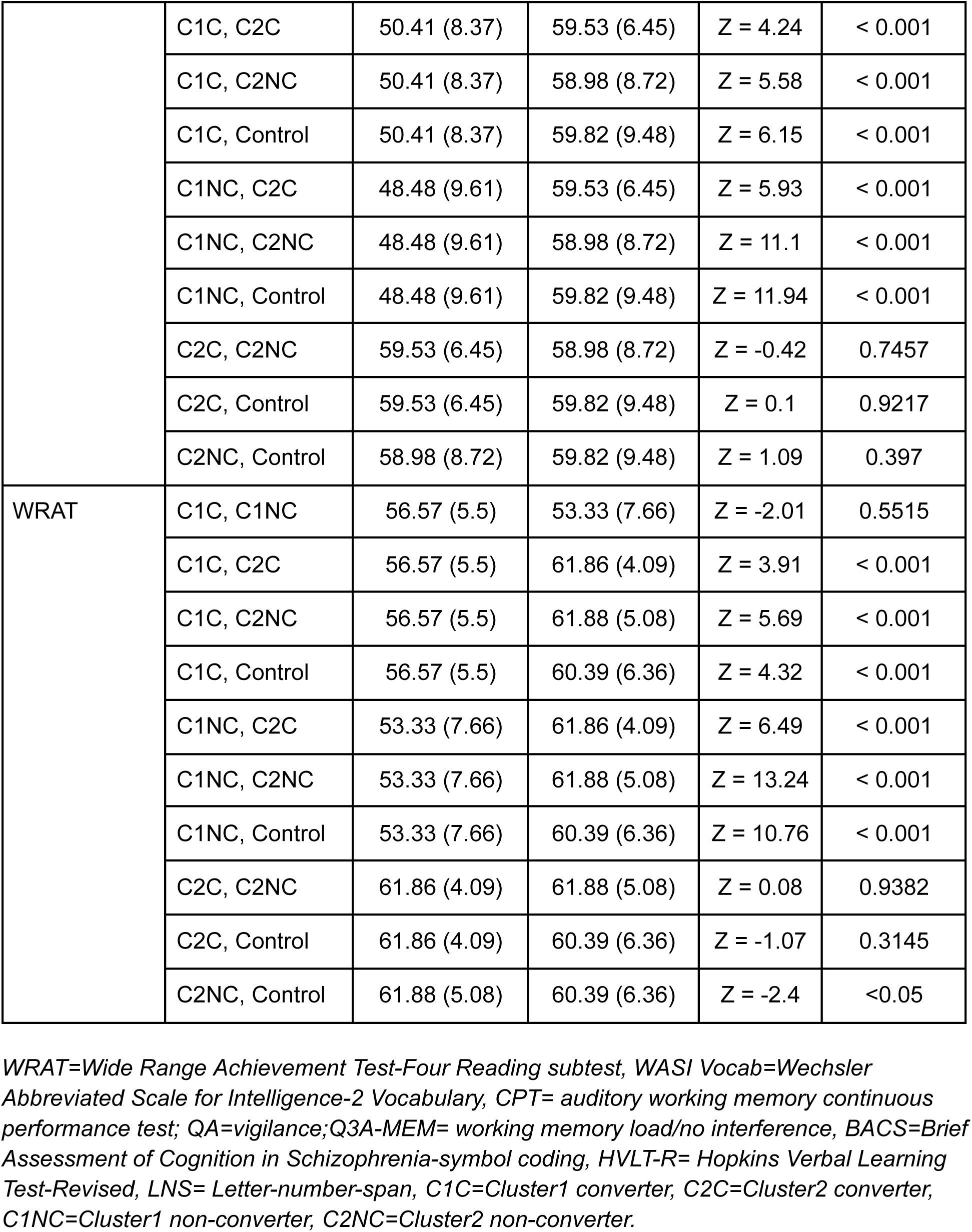
Conversion and Cluster Post-hoc Comparisons.

#### Cognition (Longitudinal)

Several cognitive variables demonstrated a “deficit” trajectory with improvement over time similar to controls. Other variables show more improvement comparatively (C1NC in WRAT (*b* = 0.57, *p* < .05), even catch up QA (*b* =1.89, *p* < 0.001) and Q3A (*b* =2.31, *p* < 0.05), and C1C in QA (*b* =3.55, *p* < 0.001), and Q3A (*b* =6.77, *p* < 0.001)). A “deterioration” trajectory was observed in C2C QA compared to the controls (*b* =-2.89, *p* < 0.05). No other cognitive variables demonstrated differences (Table 5, Fig 4).

**Figure 4:**
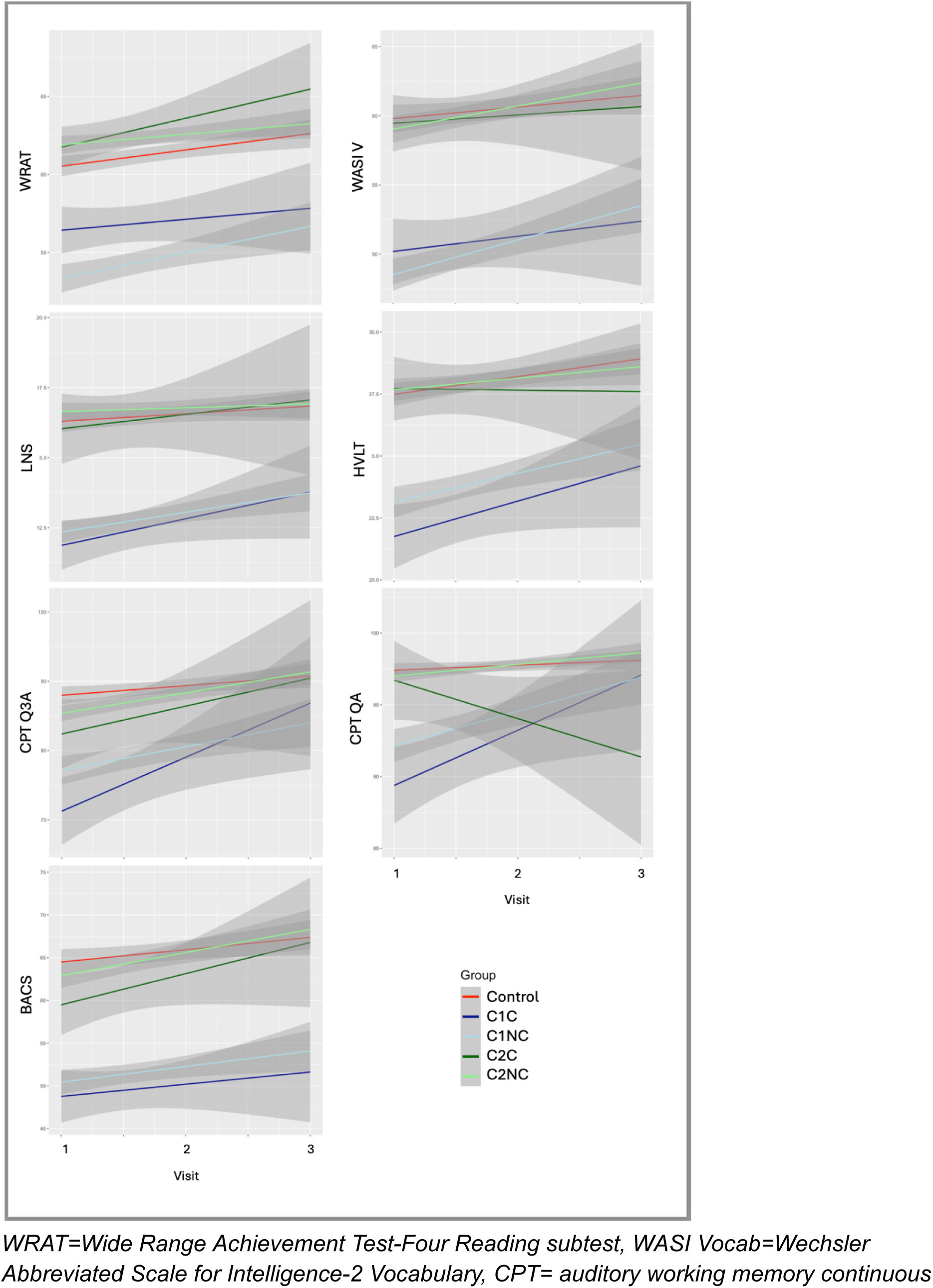
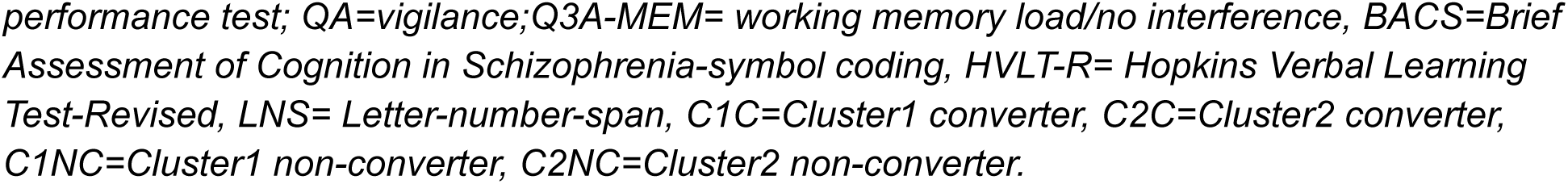
Longitudinal outcomes across cognitive variables for clusters and healthy controls.

**Table 5:** Conversion and Cluster Cognitive Longitudinal Outcomes.

|  |  | Estimate | Std.err | t | p-value |
| --- | --- | --- | --- | --- | --- |
| WRAT | (Intercept) | 59.21 | 0.46 | 129.64 | < 0.001 |
|  | C1C | -3.40 | 1.20 | -2.83 | < 0.01 |
|  | C1NC | -7.66 | 0.68 | -11.19 | < 0.001 |
|  | C2C | 0.97 | 1.45 | 0.67 | 0.5041 |
|  | C2NC | 1.71 | 0.64 | 2.66 | < 0.01 |
|  | Visit | 1.23 | 0.15 | 8.35 | < 0.001 |
|  | C1C:Visit | -0.46 | 0.48 | -0.96 | 0.423 |
|  | C1NC:Visit | 0.57 | 0.24 | 2.34 | < 0.05 |
|  | C2C:Visit | 0.40 | 0.64 | 0.63 | 0.5272 |
|  | C2NC:Visit | -0.35 | 0.22 | -1.55 | 0.1738 |
| WASI: V | (Intercept) | 58.50 | 0.69 | 85.14 | < 0.001 |
|  | C1C | -9.12 | 1.83 | -4.99 | < 0.001 |
|  | C1NC | -11.96 | 1.04 | -11.55 | < 0.001 |
|  | C2C | 1.37 | 2.20 | 0.62 | 0.5325 |
|  | C2NC | -1.12 | 0.97 | -1.16 | 0.2482 |
|  | Visit | 1.19 | 0.26 | 4.67 | < 0.001 |
|  | C1C:Visit | -0.21 | 0.82 | -0.26 | 0.7963 |
|  | C1NC:Visit | 0.83 | 0.42 | 1.95 | 0.1033 |
|  | C2C:Visit | -1.56 | 1.06 | -1.48 | 0.234 |
|  | C2NC:Visit | 0.41 | 0.39 | 1.06 | 0.361 |
| CPT: QA | (Intercept) | 97.02 | 0.64 | 151.91 | < 0.001 |
|  | C1C | -11.56 | 1.74 | -6.66 | < 0.001 |
|  | C1NC | -7.18 | 0.98 | -7.36 | < 0.001 |
|  | C2C | 2.26 | 2.12 | 1.06 | 0.2871 |
|  | C2NC | -0.81 | 0.91 | -0.89 | 0.3722 |
|  | Visit | 0.39 | 0.27 | 1.42 | 0.1559 |
|  | C1C:Visit | 3.55 | 0.86 | 4.13 | < 0.001 |
|  | C1NC:Visit | 1.89 | 0.44 | 4.31 | < 0.001 |
|  | C2C:Visit | -2.90 | 1.12 | -2.58 | < 0.05 |
|  | C2NC:Visit | 0.40 | 0.40 | 0.99 | 0.3564 |
| CPT: Q3A | (Intercept) | 86.54 | 1.24 | 69.85 | < 0.001 |
|  | C1C | -23.45 | 3.45 | -6.80 | < 0.001 |
|  | C1NC | -13.31 | 1.91 | -6.97 | < 0.001 |
|  | C2C | -7.35 | 4.21 | -1.75 | 0.0811 |
|  | C2NC | -3.97 | 1.77 | -2.24 | < 0.05 |
|  | Visit | 1.38 | 0.59 | 2.33 | < 0.05 |
|  | C1C:Visit | 6.78 | 1.91 | 3.54 | < 0.01 |
|  | C1NC:Visit | 2.32 | 0.98 | 2.37 | < 0.05 |
|  | C2C:Visit | 2.03 | 2.44 | 0.84 | 0.404 |
|  | C2NC:Visit | 1.42 | 0.89 | 1.60 | 0.123 |
| BACS | (Intercept) | 62.51 | 0.92 | 68.26 | < 0.001 |
|  | C1C | -15.43 | 2.49 | -6.20 | < 0.001 |
|  | C1NC | -13.64 | 1.39 | -9.78 | < 0.001 |
|  | C2C | -4.30 | 2.99 | -1.44 | 0.1515 |
|  | C2NC | -1.35 | 1.31 | -1.04 | 0.3006 |
|  | Visit | 1.79 | 0.40 | 4.43 | < 0.001 |
|  | C1C:Visit | 0.04 | 1.27 | 0.03 | 0.9995 |
|  | C1NC:Visit | -0.13 | 0.68 | -0.19 | 0.9999 |
|  | C2C:Visit | 0.10 | 1.62 | 0.06 | 0.9991 |
|  | C2NC:Visit | 0.00 | 0.62 | 0.00 | 0.999 |
| HVLT | (Intercept) | 26.76 | 0.38 | 70.32 | < 0.001 |
|  | C1C | -6.06 | 1.05 | -5.77 | < 0.001 |
|  | C1NC | -4.47 | 0.58 | -7.71 | < 0.001 |
|  | C2C | 0.98 | 1.27 | 0.77 | 0.4432 |
|  | C2NC | 0.63 | 0.54 | 1.16 | 0.245 |
|  | Visit | 0.69 | 0.17 | 4.04 | < 0.001 |
|  | C1C:Visit | 0.45 | 0.56 | 0.81 | 0.4924 |
|  | C1NC:Visit | 0.17 | 0.28 | 0.61 | 0.540 |
|  | C2C:Visit | -0.69 | 0.72 | -0.97 | 0.4758 |
|  | C2NC:Visit | -0.38 | 0.26 | -1.47 | 0.2817 |
| LNS | (Intercept) | 15.95 | 0.25 | 63.87 | < 0.001 |
|  | C1C | -4.58 | 0.68 | -6.74 | < 0.001 |
|  | C1NC | -4.27 | 0.38 | -11.23 | < 0.001 |
|  | C2C | 0.35 | 0.82 | 0.42 | 0.6712 |
|  | C2NC | 0.61 | 0.36 | 1.71 | 0.0872 |
|  | Visit | 0.34 | 0.11 | 3.08 | < 0.01 |
|  | C1C:Visit | 0.23 | 0.35 | 0.64 | 0.5788 |
|  | C1NC:Visit | 0.34 | 0.19 | 1.82 | 0.132 |
|  | C2C:Visit | -0.47 | 0.45 | -1.03 | 0.3778 |
|  | C2NC:Visit | -0.29 | 0.17 | -1.69 | 0.132 |

WRAT=Wide Range Achievement Test-Four Reading subtest, WASI Vocab=Wechsler Abbreviated Scale for Intelligence-2 Vocabulary, CPT= auditory working memory continuous performance test; QA=vigilance; Q3A-MEM= working memory load/no interference, BACS=Brief Assessment of Cognition in Schizophrenia-symbol coding, HVLT-R= Hopkins Verbal Learning Test-Revised, LNS= Letter-number-span, C1C=Cluster1 converter, C2C=Cluster2 converter, C1NC=Cluster1 non-converter, C2NC=Cluster2 non-converter, Std.err= Standard error.

#### Functioning (Baseline)

A significant difference in functioning was observed, where C1 showed lower GF: RS than C2 (p < 0.001), and C1 (p < 0.001), and C2 (p < 0.001), lower than controls (p < 0.001). The same trend was observed in GF: SS (C1 vs C2: p < 0.01, C1 vs HC: p < 0.001, C2 vs. HC: p < 0.001). In GAF, C1 (p < 0.001) and C2 (p < 0.001) were lower than controls. (Table 6, Fig 5).

**Figure 5:**
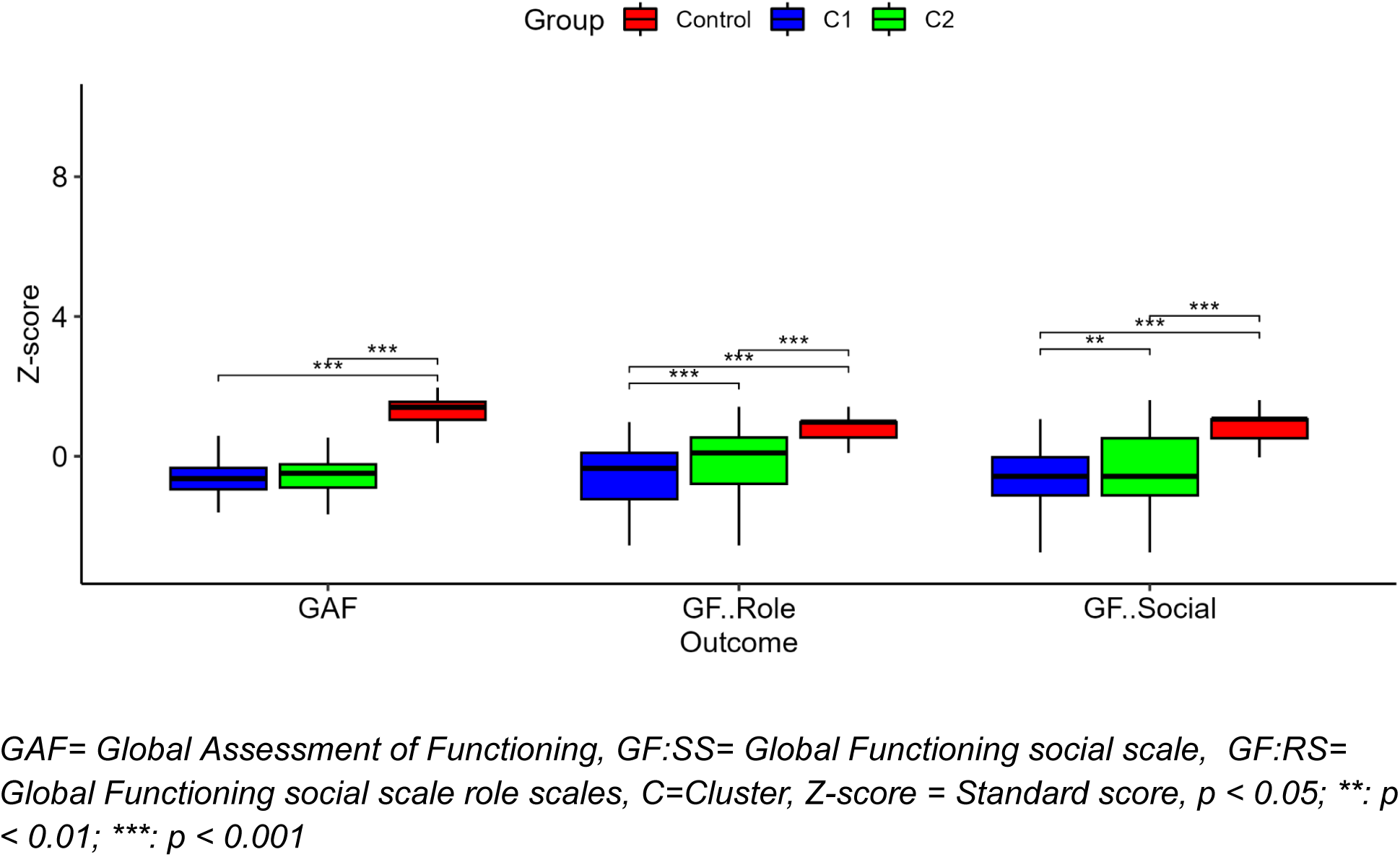
Baseline comparing in functioning between the clusters and control

**Table 6:**
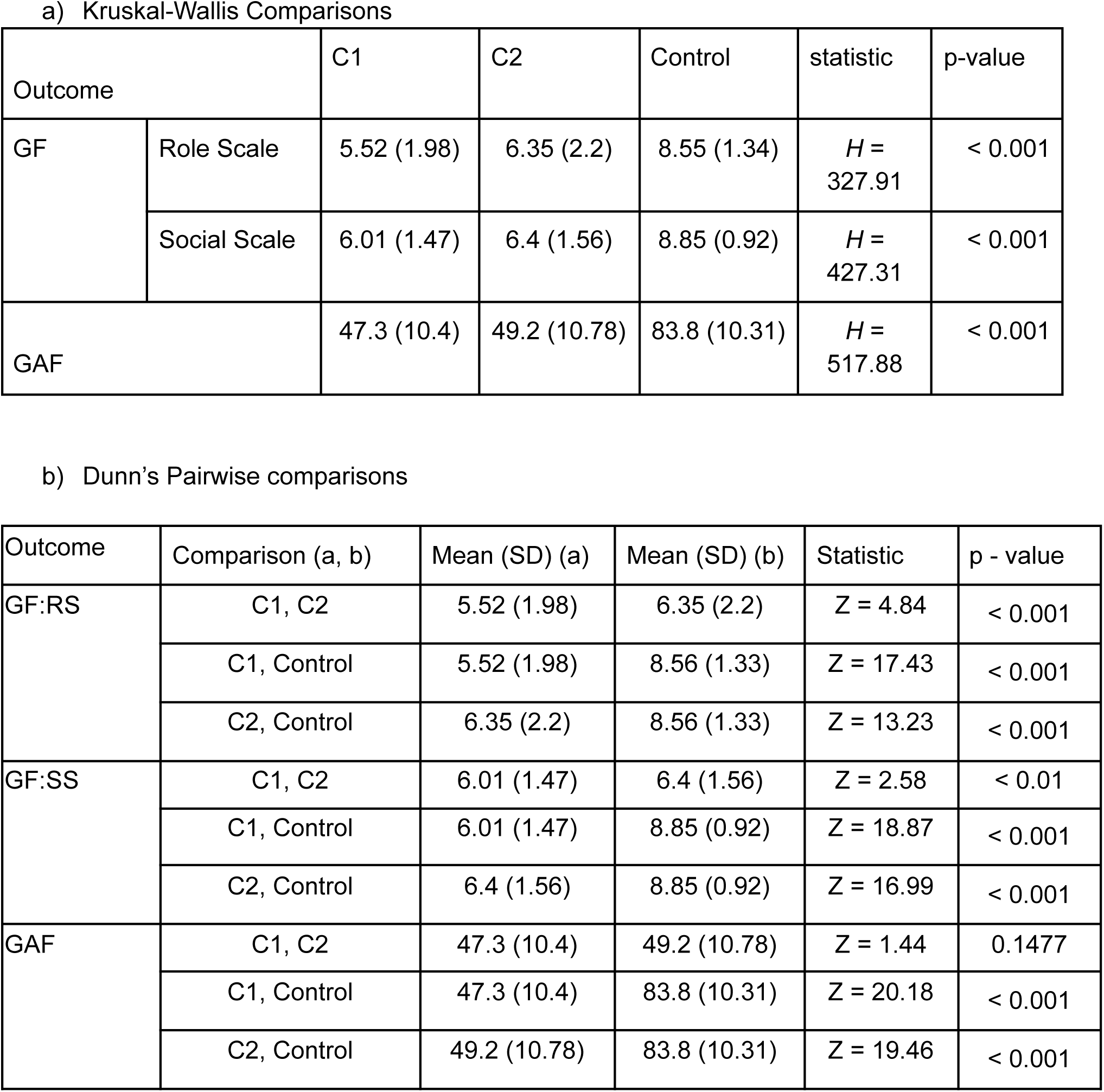
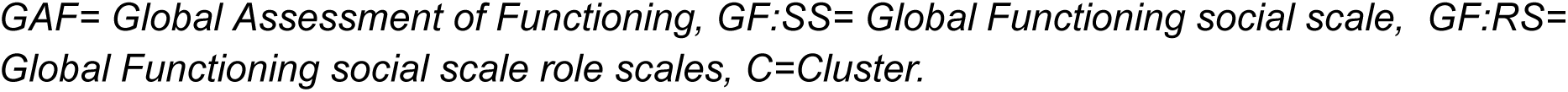
Functioning.

#### Functioning (Longitudinal)

GF: SS demonstrated a trajectory with mild improvement in both clusters (C1: (*b* = .19, *p* < 0.001), C2 (*b* = .16, *p* < 0.001)) compared to controls. This was also observed in GAF (C1: (*b* =1.74, *p* < 0.001), C2 (*b* = 1.67, *p* < 0.001)). No change was observed in GF: RS (Table 7, Fig 6).

**Figure 6:**
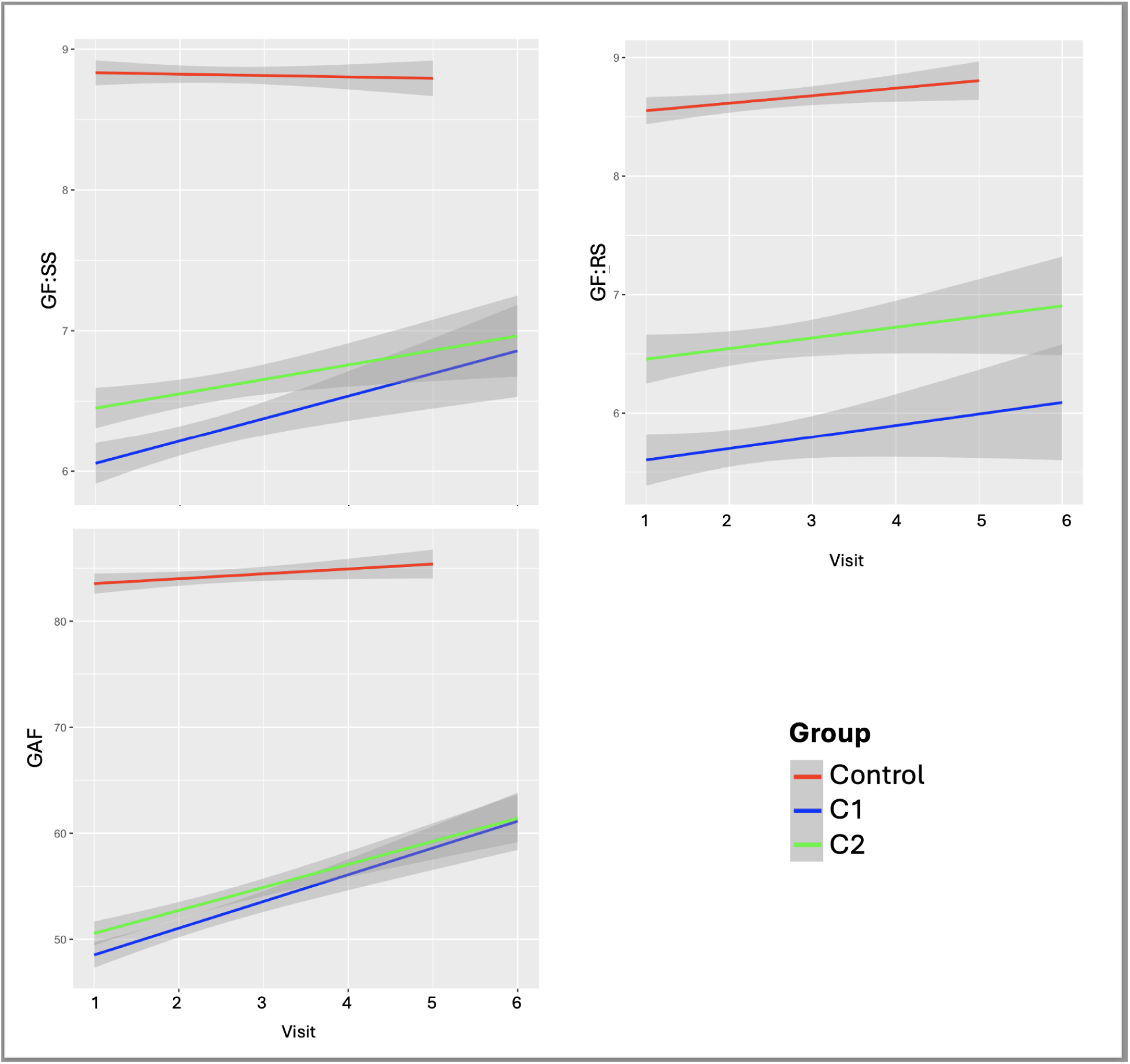
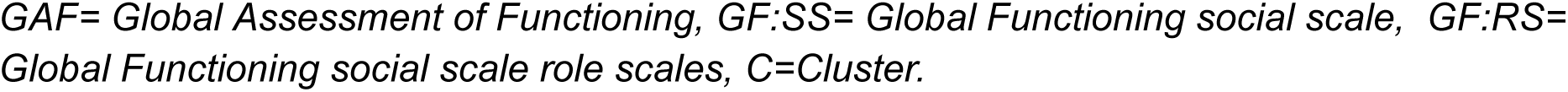
Longitudinal functioning outcomes between cluster groups and health controls.

**Table 7:** Longitudinal Outcomes for Functioning, Cluster membership, and Cluster Membership and Conversion Status.

|  |  | Estimate | Std.err | t | p-value |
| --- | --- | --- | --- | --- | --- |
| GF: RS | (intercept) | 8.51 | 0.13 | 63.77 | < 0.001 |
|  | C1 | -2.97 | 0.19 | -15.39 | < 0.001 |
|  | C2 | -2.12 | 0.18 | -11.59 | < 0.001 |
|  | Visit | 0.05 | 0.04 | 1.186 | 0.24 |
|  | C1*Visit | -0.002 | 0.06 | -0.04 | 0.97 |
|  | C2*Visit | 0.009 | 0.06 | 0.17 | 0.97 |
| GF: SS | (intercept) | 8.85 | 0.09 | 95.49 | < 0.001 |
|  | C1 | -2.97 | 0.13 | -22.30 | < 0.001 |
|  | C2 | -2.54 | 0.12 | -19.95 | < 0.001 |
|  | Visit | -0.02 | 0.03 | -0.85 | 0.399 |
|  | C1*Visit | 0.19 | 0.04 | 4.75 | < 0.001 |
|  | C2*Visit | 0.16 | 0.04 | 4.37 | < 0.001 |
| GAF | (intercept) | 83.25 | 0.75 | 111.74 | < 0.001 |
|  | C1 | -36.83 | 1.08 | -34.10 | < 0.001 |
|  | C2 | -34.83 | 1.03 | -33.96 | < 0.001 |
|  | Visit | 0.37 | 0.24 | 1.52 | 0.13 |
|  | C1*Visit | 1.74 | 0.37 | 4.58 | < 0.001 |
|  | C2*Visit | 1.67 | 0.34 | 4.86 | < 0.001 |

*GAF= Global Assessment of Functioning, GF:SS= Global Functioning social scale, GF:RS= Global Functioning social scale role scales, C=Cluster, Std.err= Standard error.*

#### Functioning (Baseline)

Pairwise comparisons demonstrated that C1C had the lowest functioning compared to C2C (p < 0.05), C2NC (p < 0.001), and controls (p < 0.001) for the GF: RS. Additionally in GF:RS, C1NC had lower functioning than C2NC (p < 0.001). In GAF, C2NC (p < 0.05) and controls (p < 0.001) had higher functioning than C1C, and C1NC (p < 0.001), C2C (p < 0.001), and C2NC (p < 0.001), lower than controls. The differences observed in GF: SS were between the controls and the cluster versus conversion groups (C1C vs. HC: p< 0.001, C1NC vs. HC: p< 0.001, C2C vs. HC: p< 0.001, C2NC vs. HC: p< 0.001) (Table 8, Fig 7).

**Figure 7:**
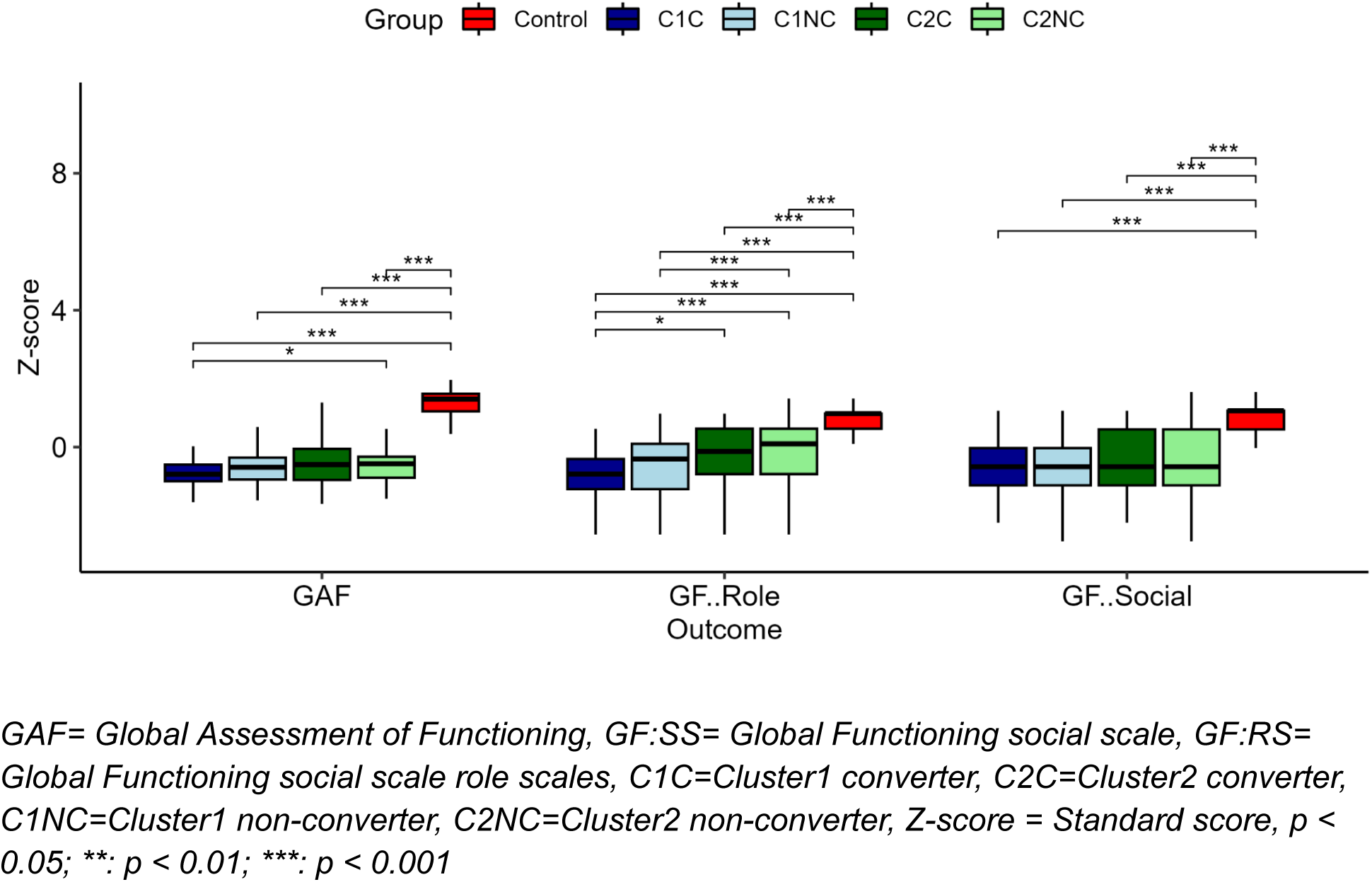
Pairwise comparisons between clusters, converter status, and healthy controls

**Table 8:** Conversion and Cluster Post-hoc Functioning Comparisons.

a) Kruskal-Wallis
| Outcome | Impaired |  | Intact |  | Control | statistic | p-value |
| --- | --- | --- | --- | --- | --- | --- | --- |
|  | Converter | NonConverter | Converter | NonConverter |  |  |  |
| GAF | 43.77<br>(9.57) | 48.1 (10.43) | 49.75<br>(12.6) | 49.13 (10.55) | 83.8<br>(10.31) | $H = 521.07$ | <0.001 |
| GF: RS | 5.08<br>(1.84) | 5.62 (2) | 6.39<br>(2.09) | 6.35 (2.22) | 8.5<br>(1.33) | $H = 330.18$ | < 0.001 |
| GF: SS | 5.84<br>(1.49) | 6.05 (1.47) | 6.42<br>(1.63) | 6.4 (1.55) | 8.85<br>(0.92) | $H = 427.71$ | < 0.001 |

b) Dunn pairwise comparisons
| Outcome | Comparison (a, b) | Mean (SD) (a) | Mean (SD) (b) | Statistic | p-value |
| --- | --- | --- | --- | --- | --- |
| GAF | C1C, C1NC | 43.76 (9.57) | 48.1 (10.43) | $Z = 1.79$ | 0.1235 |
| | C1C, C2C | 43.76 (9.57) | 49.75 (12.6) | $Z = 1.56$ | 0.16911 |
| | C1C, C2NC | 43.76 (9.57) | 49.13 (10.55) | $Z = 2.27$ | <0.05 |
| | C1C, Control | 43.76 (9.57) | 83.82 (10.31) | $Z = 12.73$ | < 0.001 |
| | C1NC, C2C | 48.1 (10.43) | 49.75 (12.6) | $Z = 0.35$ | 0.8052 |
| | C1NC, C2NC | 48.1 (10.43) | 49.13 (10.55) | $Z = 0.77$ | 0.5525 |
| | C1NC, Control | 48.1 (10.43) | 83.82 (10.31) | $Z = 18.57$ | < 0.001 |
| | C2C, C2NC | 49.75 (12.6) | 49.13 (10.55) | $Z = 0.03$ | 0.9767 |
| | C2C, Control | 49.75 (12.6) | 83.82 (10.31) | $Z = 9.03$ | < 0.001 |
| | C2NC, Control | 49.13 (10.55) | 83.82 (10.31) | $Z = 18.91$ | < 0.001 |
| GF: RS | C1C, C1NC | 5.08 (1.84) | 5.62 (2) | Z = 1.51 | 0.1461 |
|  | C1C, C2C | 5.08 (1.84) | 6.39 (2.09) | Z = 2.69 | < 0.05 |
|  | C1C, C2NC | 5.08 (1.84) | 6.35 (2.22) | Z = 3.87 | < 0.001 |
|  | C1C, Control | 5.08 (1.84) | 8.55 (1.34) | Z = 10.98 | < 0.001 |
|  | C1NC, C2C | 5.62 (2) | 6.39 (2.09) | Z = 1.95 | 0.6324 |
|  | C1NC, C2NC | 5.62 (2) | 6.35 (2.22) | Z = 3.98 | < 0.001 |
|  | C1NC, Control | 5.62 (2) | 8.55 (1.34) | Z = 16.03 | < 0.001 |
|  | C2C, C2NC | 6.39 (2.09) | 6.35 (2.22) | Z = 0.02 | 0.9838 |
|  | C2C, Control | 6.39 (2.09) | 8.55 (1.34) | Z = 6.14 | < 0.001 |
|  | C2NC, Control | 6.35 (2.22) | 8.55 (1.34) | Z = 12.85 | < 0.001 |
| GF: SS | C1C, C1NC | 5.84 (1.49) | 6.05 (1.47) | Z = 0.62 | 0.5975 |
|  | C1C, C2C | 5.84 (1.49) | 6.42 (1.63) | Z = 1.41 | 0.2245 |
|  | C1C, C2NC | 5.84 (1.49) | 6.4 (1.55) | Z = 1.89 | 0.9815 |
|  | C1C, Control | 5.84 (1.49) | 8.85 (0.92) | Z = 11.04 | < 0.001 |
|  | C1NC, C2C | 6.05 (1.47) | 6.42 (1.63) | Z = 1.18 | 0.2961 |
|  | C1NC, C2NC | 6.05 (1.47) | 6.4 (1.55) | Z = 2.15 | 0.6264 |
|  | C1NC, Control | 6.05 (1.47) | 8.85 (0.92) | Z = 17.7 | < 0.001 |
|  | C2C, C2NC | 6.42 (1.63) | 6.4 (1.55) | Z = -0.12 | 0.9071 |
|  | C2C, Control | 6.42 (1.63) | 8.85 (0.92) | Z = 7.76 | < 0.001 |
|  | C2NC, Control | 6.4 (1.55) | 8.85 (0.92) | Z = 16.54 | < 0.001 |

GAF= Global Assessment of Functioning, GF:SS= Global Functioning social scale, GF:RS= Global Functioning social scale role scales, C1C=Cluster1 converter, C2C=Cluster2 converter, C1NC=Cluster1 non-converter, C2NC=Cluster2 non-converter.

#### Functioning (Longitudinal)

In GF:SS, only C1NC (*b* = 0.22, *p* < 0.001) and C2NC (*b* = 0.17, *p* < 0.001) demonstrated a significant, but slight, improved trajectory in both clusters compared to controls. In GF:RS, C2C demonstrated significant decline overtime compared to controls (*b* = -0.57, *p* < 0.001). In GAF, C1NC (*b* = 1.97, *p* < 0.001), and C2NC (*b* = 2.09, *p* < 0.001), both demonstrated trajectories with slight improvement, while C2C demonstrated a significant decline (*b* = -3.95, *p* < 0.001). (Table 9, Figure 8).

**Figure 8:**
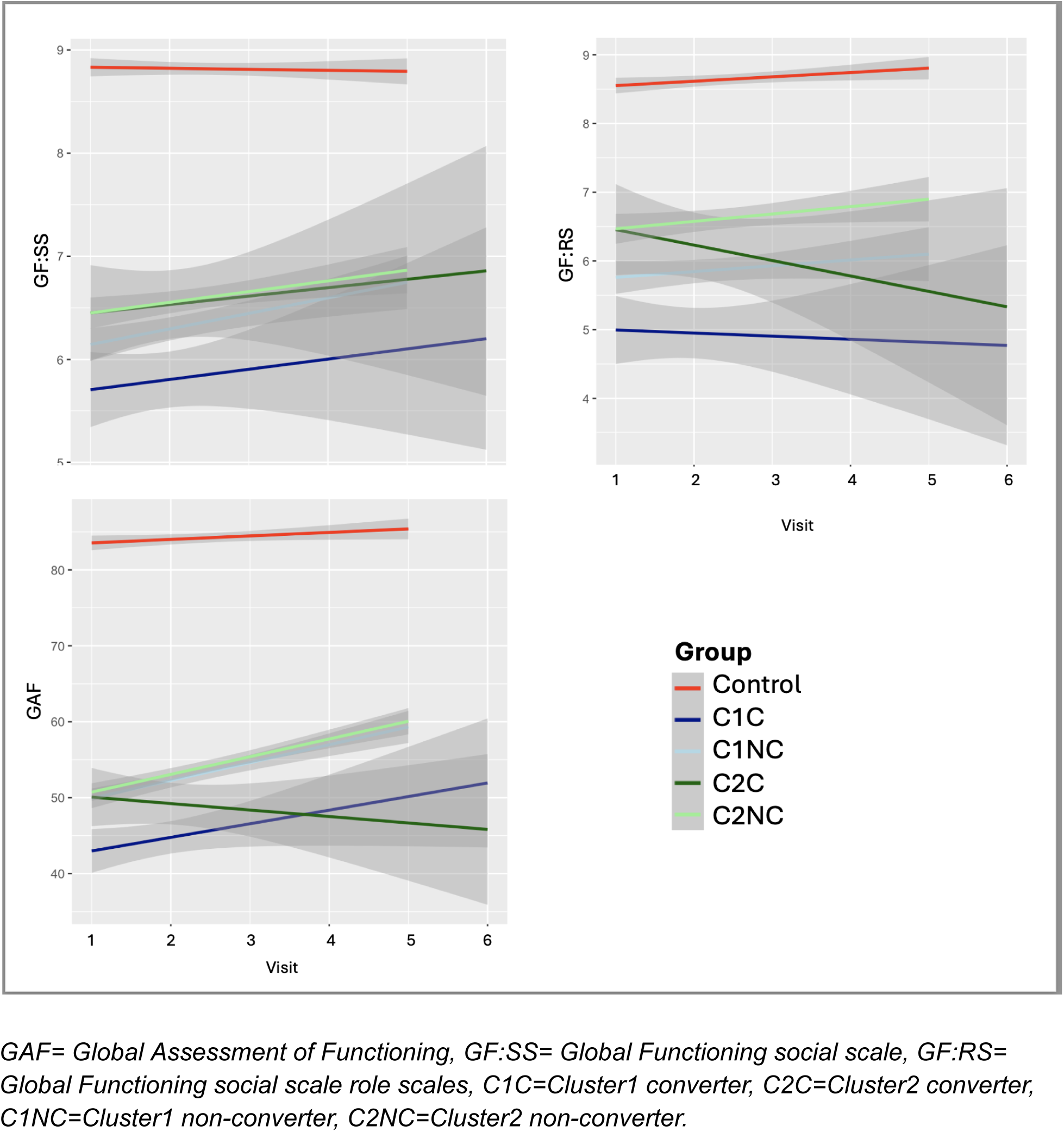
Longitudinal differences between clusters, converters, and healthy controls.

**Table 9:** Cluster and Converter Status with Functional Outcomes Longitudinal.

|  |  | Estimate | Std.err | t | p-value |
| --- | --- | --- | --- | --- | --- |
| GF: RS | (intercept) | 8.51 | 0.13 | 64.10 | <0.001 |
|  | C1C | -3.31 | 0.37 | -8.96 | <0.001 |
|  | C1NC | -2.84 | 0.20 | -14.08 | <0.001 |
|  | C2C | -1.39 | 0.42 | -3.32 | <0.001 |
|  | C2NC | -2.17 | 0.19 | -11.59 | <0.001 |
|  | Visit | 0.05 | 0.04 | 1.20 | 0.23 |
|  | C1C*Visit | -0.25 | 0.15 | -1.73 | 0.12 |
|  | C1NC*Visit | 0.02 | 0.06 | 0.33 | 0.74 |
|  | C2C*Visit | -0.57 | 0.15 | -3.73 | <0.001 |
|  | C2NC*Visit | 0.05 | 0.06 | 0.93 | 0.394 |
| GF: SS | (intercept) | 8.85 | 0.09 | 95.63 | <0.001 |
|  | C1C | -2.98 | 0.25 | -11.77 | <0.001 |
|  | C1NC | -2.93 | 0.14 | -20.91 | <0.001 |
|  | C2C | -2.34 | 0.29 | -8.11 | <0.001 |
|  | C2NC | -2.55 | 0.13 | -19.56 | <0.001 |
|  | Visit | -0.02 | 0.03 | -0.85 | 0.40 |
|  | C1C*Visit | -0.08 | 0.09 | -0.82 | 0.41 |
|  | C1NC*Visit | 0.22 | 0.04 | 5.32 | <0.01 |
|  | C2C*Visit | 0.01 | 0.10 | 0.10 | 0.92 |
|  | C2NC*Visit | 0.17 | 0.04 | 4.61 | <0.001 |
| GAF | (intercept) | 83.25 | 0.73 | 113.29 | <0.001 |
|  | C1C | -39.41 | 2.10 | -18.81 | <0.001 |
|  | C1NC | -35.81 | 1.12 | -32.01 | <0.001 |
|  | C2C | -28.36 | 2.35 | -12.06 | <0.001 |
|  | C2NC | -35.19 | 1.04 | -33.89 | <0.001 |
|  | Visit | 0.37 | 0.24 | 1.55 | 0.12 |
|  | C1C*Visit | -0.59 | 0.89 | -0.65 | 0.513 |
|  | C1NC*Visit | 1.97 | 0.39 | 5.10 | <0.001 |
|  | C2C*Visit | -3.95 | 0.93 | -4.26 | <0.001 |
|  | C2NC*Visit | 2.09 | 0.34 | 6.06 | <0.001 |

GAF= Global Assessment of Functioning, GF:SS= Global Functioning social scale, GF:RS= Global Functioning social scale role scales, C1C=Cluster1 converter, C2C=Cluster2 converter, C1NC=Cluster1 non-converter, C2NC=Cluster2 non-converter.

#### Clinical (Baseline)

The clusters did not demonstrate baseline differences in SOPS positive, negative, disorganization, or general domains (Table 10, Fig. 9).

**Figure 9:**
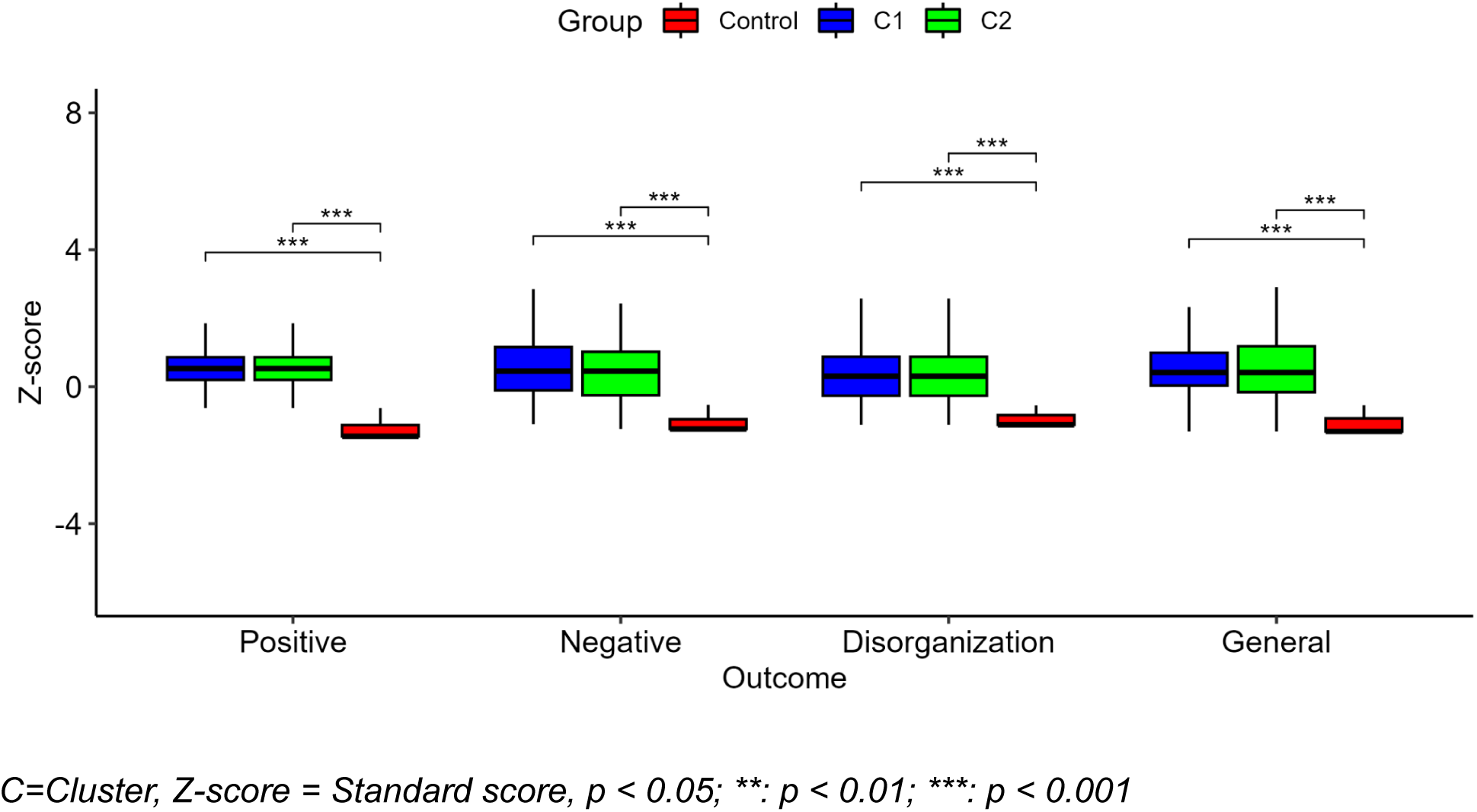
Differences in the Scale of Prodromal Symptoms between the clusters and control

**Table 10:** Clinical and Cognitive Cluster pairwise comparisons.

a) Kruskal-Wallis Comparisons
| Outcome |  | C1 | C2 | Control | statistic | p-value |
| --- | --- | --- | --- | --- | --- | --- |
| SOPS | Positive | 12.21 (3.7) | 12.18 (3.68) | 1.07 (1.67) | $H = 563.44$ | $< 0.001$ |
| | Negative | 12.46 (5.96) | 11.6 (6.09) | 1.46 (2.23) | $H = 482.28$ | $< 0.001$ |
| | Disorganization | 5.55 (3.36) | 5.21 (3.12) | 0.66 (1.19) | $H = 451.72$ | $< 0.001$ |
| | General | 9.37 (4.11) | 9.19 (4.33) | 1.34 (2.16) | $H = 462.51$ | $< 0.001$ |

b) Dunn's Pairwise comparisons
| Outcome | Comparison (a, b) | Mean (SD) (a) | Mean (SD) (b) | Statistic | p-value |
| --- | --- | --- | --- | --- | --- |
| Positive | C1, C2 | 12.21 (3.7) | 12.18 (3.68) | $Z = 0$ | 0.9963 |
| | C1, Control | 12.21 (3.7) | 1.06 (1.67) | $Z = -19.99$ | $< 0.001$ |
| | C2, Control | 12.18 (3.68) | 1.06 (1.67) | $Z = -20.71$ | $< 0.001$ |
| Negative | C1, C2 | 12.46 (5.96) | 11.6 (6.09) | $Z = -1.37$ | 0.1715 |
| | C1, Control | 12.46 (5.96) | 1.49 (2.28) | $Z = -19.11$ | $< 0.001$ |
| | C2, Control | 11.6 (6.09) | 1.49 (2.28) | $Z = -18.45$ | $< 0.001$ |
| Disorganization | C1, C2 | 5.55 (3.36) | 5.21 (3.12) | $Z = -0.82$ | 0.4131 |
| | C1, Control | 5.55 (3.36) | 0.64 (1.18) | $Z = -18.35$ | $< 0.001$ |
| | C2, Control | 5.21 (3.12) | 0.64 (1.18) | $Z = -18.19$ | $< 0.001$ |
| General | C1, C2 | 9.37 (4.11) | 9.19 (4.33) | $Z = -0.47$ | 0.6379 |
| | C1, Control | 9.37 (4.11) | 1.3 (2.12) | $Z = -18.4$ | $< 0.001$ |
| | C2, Control | 9.19 (4.33) | 1.3 (2.12) | $Z = -18.62$ | $< 0.001$ |

SD= Standard deviation, C=Cluster

#### Clinical (Longitudinal)

For Positive symptoms, converter groups demonstrated worsening symptom trajectories (C1C: *b* = 3.53, *p* < 0.001; C2C: *b* = 2.27, *p* <0.001) while non-converter groups showed improvement trajectories (C1NC: *b* = -2.26, *p* <0.001; C2NC: *b* = -2.04, *p* <0.001) compared to controls. In Negative, Disorganization, and General symptoms non-converter groups demonstrated significant improvement compared to controls (Negative (C1NC: *b* = -1.89, *p* <0.001; C2NC: *b* = -1.95, *p* <0.001); Disorganization (C1NC: *b* = -0.81, *p* <0.0001; C2NC: *b* = -0.92, *p* <0.0001); General(C1NC: *b* = -1.43, *p* <0.0001; C2NC: *b* = -1.50, *p* <0.0001), while converter groups didn’t change significantly over time. (Table S4-S5, Fig. S2-S3).

#### Time to conversion

The Kaplan-Meyer survival curve and log-rank test demonstrated that individuals in C1 have a lower survival probability than C2 that is consistent over time *(X*(1) *=* 8.4, *p* < 0.01) (Fig. S4.a). For the sample having participants who remained in the study for < 2.5 years, C1 continued to have significantly lower survival than C2 consistently (*X*(1) = 8.1, *p* < 0.01) (Fig. S4.b).

#### Converter vs. non-convert proportion

The converters constitute 18% of C1 and 11% of C2 (*X*^2^=6, p=0.02).

#### Relationship between cognition and SOPS

The GEE showed that only negative symptoms and disorganization are associated with cognition in the non-converters, while positive symptoms are not associated with any of the cognitive measures in both converters and non converters (Table S6).

#### Relationship between cognition and functioning

The GEE showed that all the cognitive variables have a significant and positive relationship with functioning (Table S7, S8). Variables related to attention and working memory, verbal abilities, and declarative memory, had the largest estimates.

#### Converter diagnosis

In C1C, 81.9%, and in C2C 62%, had a diagnosis code that falls within the psychotic disorder diagnoses (*X*^2^=5, p=0.02). In contrast, C1C had 16.8% and C2C had 44.7% bipolar diagnosis (*X*^2^=7, p=0.006).

## Discussion

To our knowledge, this is the first and largest study to use machine learning clustering on cognitive variables in a CHR sample, independently validate the clusters with an equally large CHR sample having common cognitive variables, and investigate the clusters within the conversion status. Our findings reveal two CHR cognitive subtypes with “impaired” and “intact” cognition compared to controls. Previous work by Haining et al. and Allott et al. reported on two cognitive clusters with “intact” and “impaired” cognitive profiles as well (25,27). This is also consistent more broadly with results from the psychosis spectrum (23,66–69). While other studies report varying numbers of clusters, these differences could be attributed to the inclusion of different conditions or a mix of variable types beyond cognition in the clustering (70).

Notably, only attention and working memory as well as verbal abilities domains demonstrated differences between the cognitively intact CHR group, specifically non-converters, and controls at baseline. Despite that group having comparable cognitive abilities to controls in most domains, independent of the conversion status, it is still unable to perform as well on CPT Q3A. This task can be difficult, even for the controls, and thus assessments challenging attention and working memory abilities could potentially be an important cognitive marker to distinguish between the cognitively intact CHR group and controls early on. Our previous work does demonstrate worse performance by the CHR group, compared to controls, on attention and working memory abilities than any of the other domains, which is consistent with the current findings (35).

Verbal abilities at baseline, on the other hand, were higher in the CHR group. DeRosse et al. also reported significantly higher WRAT scores in the psychotic disorder group, which they have attributed to a compensation mechanism or “resilient factor against clinically significant psychosis” indicated by negative symptoms (71). We do not observe differences in negative symptoms, or any other SOPS subscores, between the cognitively intact and impaired CHR groups at baseline, nor do we observe a longitudinal association between WRAT and negative symptoms in the CHR group as a whole (71). While the converters had worsened positive symptoms overtime, which is expected after conversion, the non converters improved across all SOPS subscores. The association we found between negative symptoms and most of the cognitive assessments, especially CPT QA, is in alignment with previous studies. Nonetheless, others, e.g., Allott et al., did not observe an association between cognition and symptomatology, even longitudinally (27,72). This could still be a compensation mechanism, but not necessarily specifically attributed to differences in psychosis related clinical findings in the converter group, at least those measured by SOPS, in our study. Rather our results are more in alignment with Walter et al. demonstrating that, independent of symptomatology, verbal abilities are more resistant to the impact of psychiatric conditions, specifically schizophrenia, than the rest of the cognitive domains that can be quite vulnerable (73). So it is possible that the cognitive domains, other than verbal abilities, that are comparable between the cognitively intact CHR group and the controls at the time of assessment could have even been higher if it weren’t for their CHR status.

The attention and working memory domains follow a “catch up” trajectory in the cognitively impaired CHR group, independent of the conversion status, with QA reaching closer to the controls than Q3A, probably due to the task’s difficulty. In the theoretical framework by Allott et al., they describe the “deficit” trajectory in which there is no change in the trajectory itself, but the deficit is consistent throughout the 12 month follow-up from baseline. All cognitive trajectories in the cognitively impaired CHR group in our study follow this pattern except for attention and working memory domains (CPT QA and Q3A) and less so in verbal abilities (WRAT). Thus, our results support those of Allott et al. and extend their findings up to 2 years (27). The impaired cognition groups would benefit from early intervention by boosting their catch up trajectory to meet the performance of the controls.

The cognitively intact CHR group on the other hand did not demonstrate any significant trajectory differences from controls, probably because of a ceiling effect. Interestingly, however, the baseline differences in CPT Q3A distinguishing between the cognitively intact CHR group and controls fades away after 24 months with a gradual, non-significant, “catch up” trajectory. It is possible that the increased vigilance we see in the CHR post conversion, where they are more wary of their surroundings, could have contributed to this sort of compensation mechanism. This can be more clearly captured in the cognitively impaired CHR group, due to the lack of a ceiling effect, with improvement beyond the practice effects seen in the controls.

Further examination of these trajectories showed that the attention and working memory domain in the cognitively intact converter group demonstrates a sharp decline. The deterioration in CPT QA, after conversion, without an associated decline in CPT Q3A might indicate that attention, or vigilance, is more impacted than working memory. Given the substantial decline in this group, we believe that they might require close observation. Since this group starts seemingly similar to controls, clinicians might assume that they are not in need of therapeutic intervention, which might be a missed opportunity for improvement of outcome or at least sustaining their pre conversion capacity. Therapeutic interventions targeting vigilance might be most useful for this group (74).

Unlike GAF, the role and social scales were designed to assess functioning specifically in the CHR population (75), and thus it was unsurprising to observe differences in the global functioning role and social scales between the cognitively intact and impaired CHR groups, but not in GAF (75). After 24 months, however, the social scale becomes indistinguishable between the two CHR groups. More detailed examination of functioning demonstrated that the cognitively intact converters drop drastically in both GAF and role scale after conversion. This resembles the deterioration observed in CPT QA in that group as well. Our results do indicate a positive relationship between CPT QA and functioning, however, the point change is minimal compared to the association with other cognitive domains when the response variable is role scale and the predictor is CPT QA. On the other hand, when these are reversed, role scale is predictor and CPT QA is response, the estimate increases substantially. Meaning that the change in functioning has more impact on the change in cognition here than cognition has on functioning. Nonetheless, we do acknowledge the trajectory heterogeneity within CPT QA in the cognitively intact group, and thus an approach similar to Allott et al. (27), investigating CPT QA, is warranted. It is also possible that there are mediators informing the association between CPT QA and functioning that would be worth exploring.

The CHR group with impaired cognition has a significantly higher number of converters and the members convert earlier overall than the cognitively intact group. This is supported by previous research in CHR and the schizophrenia spectrum, demonstrating that neurocognitive performance at baseline is generally lower in those who later convert (76). The converters with impaired cognition also had a significantly higher percentage of a psychotic disorder diagnosis than the intact groups and significantly lower bipolar disorder diagnosis (23,24). It is possible that those who convert and have impaired cognition will end up converting to schizophrenia, which makes cognition be an important predictor of future diagnosis. This is consistent with prior work indicating that cognitive impairment of those with schizophrenia exists early in life preceding onset and is more pronounced, while those with bipolar develop such impairment later in adulthood but not as severe (23,24,77).

There is a high potential for tailored and preventative cognitive therapeutic interventions here. For example, It is intuitive that the cognitively impaired group would be recommended for cognitive intervention given their cognitive abilities early on. Therapeutic approaches here might improve their catch-up trajectory to reach a level similar to controls. Those with intact cognition on the other hand, even though they are similar to controls at baseline and follow up in most of the measures, those of them who convert, change drastically in both vigilance, and functioning, especially role scale. This group should be under close observation, as they initially seem that they would not benefit from cognitive intervention, but they actually need it to prevent their deterioration later on (38,39). Unlike what has been reported previously– that those with good cognition at baseline might not benefit from cognitive therapeutic intervention (27), we recommend that this is taken with more consideration and scrutiny to better understand those with intact cognition who end up converting.

## Conclusion

This study identified impaired and intact cognitive subtypes, independent of their conversion status. The results further indicate that attention and working memory are important to distinguish between those who are CHR with intact cognition and controls. The cognitively intact CHR group becomes less vigilant generally post conversion after 24 month followup, while the cognitively impaired one demonstrates a catch up trajectory on both attention and working memory. Overall, early assessment, covering several cognitive domains, especially attention and working memory, is crucial for identifying trajectories of improvement and deterioration for the purpose of tailoring intervention for improving outcomes in individuals at CHR.

### Limitations

There are some limitations to consider: 1) Like other longitudinal studies, the drop out rate of the participants is a limitation. 2) This is a secondary data analysis study, meaning that we were limited by the variables available to us. Additionally, the cognitive measures were limited by what is common between the discovery and validation dataset. Nonetheless, the variables included in this study do explore a wide range of cognitive domains. 3) The causality, and its direction, between the observed decline in cognition and functioning could not be established in this current work.

## Data Availability

All data are available online at the National Institute of Mental Health Data Archive.

https://nda.nih.gov/

## Acknowledgments

This work was supported by the National Institute of Mental Health (R21MH133001 to Dr Yassin; U01MH081984 to Dr Addington; R01MH60720, U01MH081944 and K24MH76191 to Dr Cadenhead; P50MH066286 (Prodromal Core) to Dr Bearden; U01MH082004 to Dr Perkins; U01MH081988 to Dr Walker; U01MH082022 to Drs Cadenhead and Woods; U01MH081928 to Dr Stone; U01MH081902 to Drs Bearden andCannon; U01MH076989 to Dr Mathalon; and U01MH081857 to Dr Cornblatt).

