## Supplementary materials for "Cognitive subtypes in youth at clinical high risk for psychosis"

**Table S1:** Comparison of NAPLS2 and NAPLS3 demographics between CHR and control participants:

| | NAPLS2 (N = 1044) | | | | NAPLS3 (N = 712) | | | |
|---|---|---|---|---|---|---|---|---|
| | Mean (SD) or N (%) | | Statistics | p value | Mean (SD) or N (%) | | Statistics | p value |
| | Control (N=280) | CHR (N=764) | | | Control (N=84) | CHR (N=628) | | |
| Age | 19.73 (4.67) | 18.50 (4.23) | $t$ = 3.85 | < 0.001 | 18.63 (4.26) | 18.25 (4.09) | $t$ = 0.76 | 0.44 |
| Sex, male: female | 141 (50.4%) | 436 (57.1%) | $X^2$ = 3.47 | 0.06 | 41 (48.8%) | 342 (54.5%) | $X^2$ = 0.73 | 0.39 |
| Income | | | $X^2$ = 12.49 | 0.05 | | | $X^2$ = 7.99 | 0.239 |
| Less than 10,000 | 23 (8.2%) | 72 (9.5%) | | | 9 (10.7%) | 52 (8.3%) | | |
| 10,000 – 19,999 | 20 (7.2%) | 73 (9.7%) | | | 10 (11.9%) | 39 (6.3%) | | |
| 20,000 – 39,999 | 37 (13.3%) | 86 (11.4%) | | | 9 (10.7%) | 57 (9.1%) | | |
| 40,000 – 59,999 | 33 (11.8%) | 78 (10.3%) | | | 8 (9.5%) | 58 (9.3%) | | |
| 60,000 – 99,999 | 55 (19.7%) | 106 (14.0%) | | | 16 (19.0%) | 92 (14.8%) | | |
| 100,000 + | 68 (24.4%) | 170 (22.5%) | | | 18 (21.4%) | 171 (27.4%) | | |
| Don't know or refused | 43 (15.4%) | 171 (22.6%) | | | 14 (16.7%) | 154 (24.7%) | | |
| Education | | | $X^2$ = 89.83 | < 0.001 | | | $X^2$ = 17.46 | < 0.01 |
| No High School | 101 (36.1%) | 400 (52.8%) | | | 35 (41.7%) | 326 (51.9%) | | |
| High School | 92 (32.9%) | 289 (38.1%) | | | 30 (35.7%) | 235 (37.4%) | | |

| | | | | | |
|---|---|---|---|---|---|
| College | 25 (8.9%) | 23 (3.0%) | | 4 (4.8%) | 29 (4.6%) |
| Technical school | 4 (1.4%) | 12 (1.6%) | | 2 (2.4%) | 2 (0.3%) |
| University | 47 (16.8%) | 29 (3.8%) | | 11 (13.1%) | 31 (4.9%) |
| Graduate school | 11 (3.9%) | 5 (0.7%) | | 2 (2.4%) | 5 (0.8%) |

*CHR= Clinical high risk for psychosis, SD = standard deviation, N=sample size*

**Table S2:** Silhouette scores

| | Silhouette | | | | | | | | |
|---|---|---|---|---|---|---|---|---|---|
| Cluster number | 2 | 3 | 4 | 5 | 6 | 7 | 8 | 9 | 10 |
| k means | 0.4288 | 0.2004 | 0.2232 | 0.1967 | 0.2084 | 0.1002 | 0.0484 | 0.1002 | 0.0718 |
| Agnes | 0.4434 | 0.3447 | 0.2581 | 0.2700 | 0.2814 | 0.2419 | 0.1799 | 0.2200 | 0.1908 |
| Diana | 0.5045 | 0.4072 | 0.3532 | 0.3105 | 0.2993 | 0.2916 | 0.2916 | 0.2927 | 0.2748 |
| PAM | 0.5028 | 0.3990 | 0.3593 | 0.3646 | 0.3710 | 0.3956 | 0.3794 | 0.3551 | 0.3486 |

PAM=Partition Around Medoids, Agnes=AGglomerative NESting, and DIANA=DIvisive ANAlysis Clustering. Parameters include, nClust num= 2; metric=correlation, method= average, neighbor size= 3.

**Table S3:** Confusion Matrix in validation sample (NAPLS3)

| | | Actual Values | |
|---|---|---|---|
| | | C1 | C2 |
| Predicted Values | C1 | 378 | 64 |
| | C2 | 50 | 199 |

Accuracy: 83%, Precision: 88%, Recall 86%, F1-Score 87%, C= Cluster

**Figure S1:** Cluster 3D Visualization and Dendrograms

a) Discovery Sample (NAPLS2)

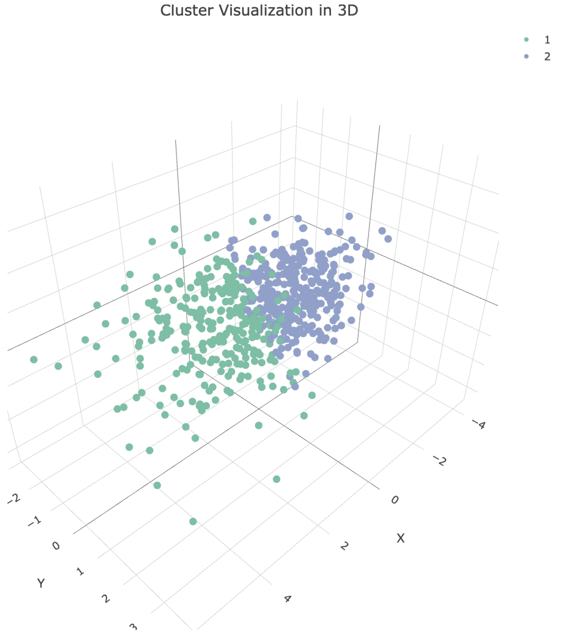

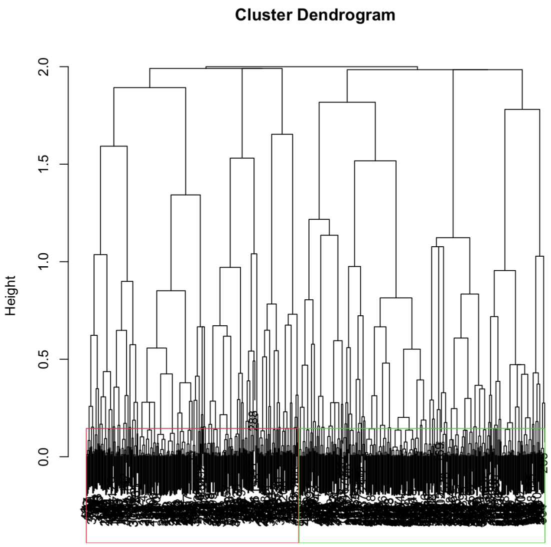

### b) Validation sample (NAPLS3)

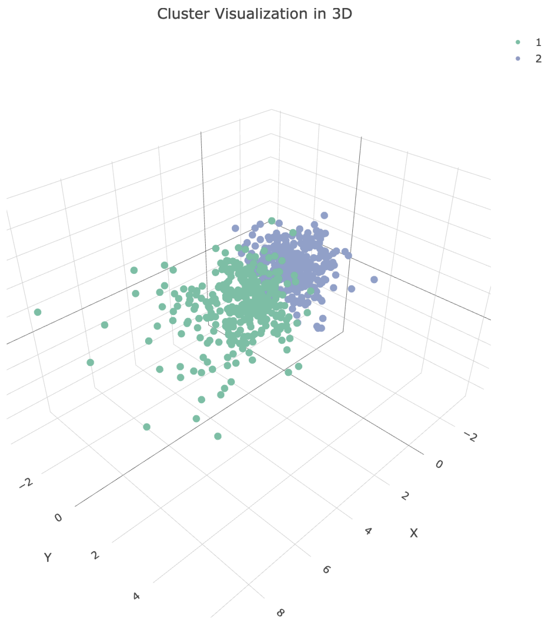

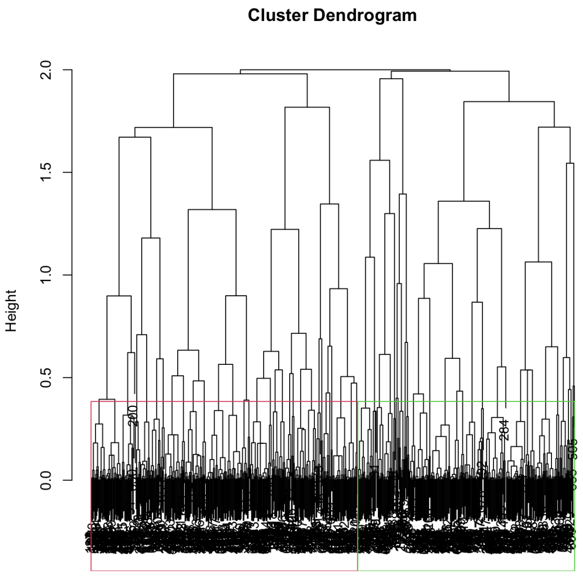

**Table S4:** Longitudinal Results for Clusters and SIPS

| | | Estimate | Std.err | t | p-value |
|---|---|---|---|---|---|
| Positive | (Intercept) | 1.24 | 0.30 | 4.12 | < 0.0001 |
| | C1 | 12.32 | 0.45 | 27.54 | < 0.0001 |
| | C2 | 12.77 | 0.43 | 29.98 | < 0.0001 |
| | Visit | -0.22 | 0.19 | -1.18 | 0.2396 |
| | C1*Visit | -1.21 | 0.30 | -4.10 | < 0.0001 |
| | C2*Visit | -1.69 | 0.28 | -6.09 | < 0.0001 |
| Negative | (Intercept) | 1.55 | 0.44 | 3.52 | < 0.001 |
| | C1 | 12.38 | 0.64 | 19.34 | < 0.0001 |
| | C2 | 11.73 | 0.61 | 19.14 | < 0.0001 |
| | Visit | -0.11 | 0.21 | -0.53 | 0.5992 |
| | C1*Visit | -1.50 | 0.33 | -4.56 | < 0.0001 |
| | C2*Visit | -1.79 | 0.31 | -5.79 | < 0.0001 |
| Disorganization | (Intercept) | 0.61 | 0.23 | 2.62 | < 0.05 |
| | C1 | 5.46 | 0.34 | 16.14 | < 0.0001 |
| | C2 | 5.38 | 0.32 | 16.59 | < 0.0001 |
| | Visit | 0.01 | 0.11 | 0.12 | 0.9079 |
| | C1*Visit | -0.58 | 0.17 | -3.37 | < 0.01 |
| | C2*Visit | -0.86 | 0.16 | -5.31 | < 0.0001 |
| General | (Intercept) | 1.48 | 0.33 | 4.45 | < 0.0001 |
| | C1 | 9.15 | 0.48 | 18.87 | < 0.0001 |
| | C2 | 9.09 | 0.46 | 19.59 | < 0.0001 |
| | Visit | -0.16 | 0.17 | -0.97 | 0.3351 |
| | C1*Visit | -1.26 | 0.26 | -4.85 | < 0.0001 |
| | C2*Visit | -1.37 | 0.24 | -5.62 | < 0.0001 |

*Positive=SIPS: positive symptom sum score, Negative=SIPS: negative symptom sum score, Disorganization= SIPS disorganization symptom sum score, General=SIPS general symptom sum score, C=Cluster, Std.err= Standard error.*

**Figure S2:** Longitudinal differences between clusters, and healthy controls.

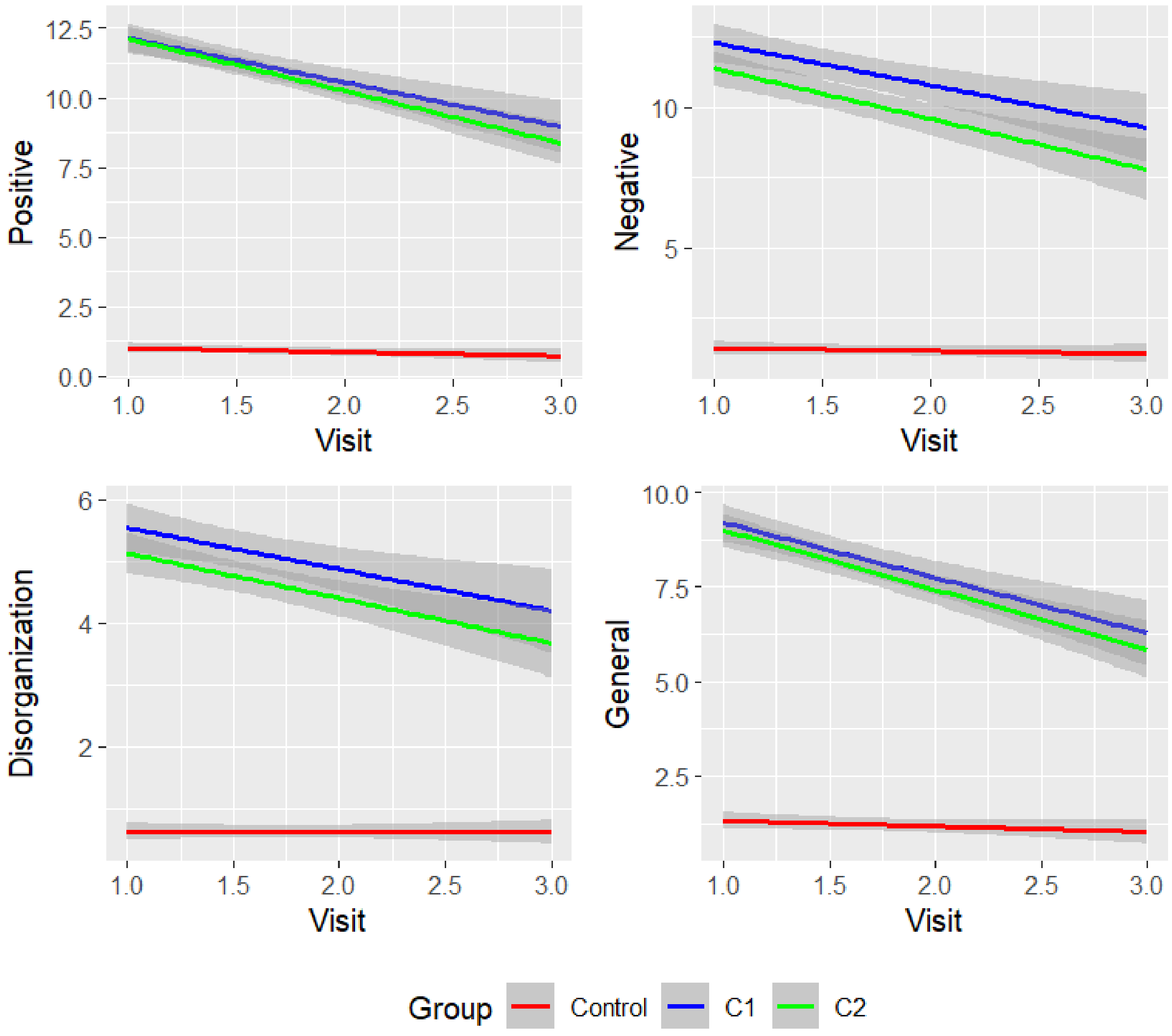

**Table S5:** Longitudinal results for Clusters, Converter Status, and SIPS.

| | | Estimate | Std.err | t | p - value |
|---|---|---|---|---|---|
| Positive | (Intercept) | 1.23 | 0.29 | 4.27 | < 0.0001 |
| | C1C | 8.68 | 0.82 | 10.54 | < 0.0001 |
| | C1NC | 13.01 | 0.45 | 29.20 | < 0.0001 |
| | C2C | 10.59 | 1.00 | 10.59 | < 0.0001 |
| | C2NC | 12.85 | 0.41 | 31.01 | < 0.0001 |
| | Visit | -0.21 | 0.16 | -1.27 | 0.2057 |
| | C1C*Visit | 3.53 | 0.52 | 6.79 | < 0.0001 |
| | C1NC*Visit | -2.26 | 0.27 | -8.21 | < 0.0001 |
| | C2C*Visit | 2.27 | 0.66 | 3.46 | < 0.001 |
| | C2NC*Visit | -2.04 | 0.25 | -8.19 | < 0.0001 |
| Negative | (Intercept) | 1.55 | 0.44 | 3.54 | < 0.001 |
| | C1C | 11.18 | 1.21 | 9.23 | < 0.0001 |
| | C1NC | 12.52 | 0.67 | 18.69 | < 0.0001 |
| | C2C | 9.42 | 1.47 | 6.41 | < 0.0001 |
| | C2NC | 11.90 | 0.63 | 19.03 | < 0.0001 |
| | Visit | -0.11 | 0.21 | -0.53 | 0.6607 |
| | C1C*Visit | 0.62 | 0.66 | 0.94 | 0.4327 |
| | C1NC*Visit | -1.89 | 0.34 | -5.57 | < 0.0001 |
| | C2C*Visit | 0.28 | 0.84 | 0.33 | 0.7383 |
| | C2NC*Visit | -1.95 | 0.31 | -6.28 | < 0.0001 |
| Disorganization | (Intercept) | 0.61 | 0.23 | 2.62 | < 0.05 |
| | C1C | 6.42 | 0.63 | 10.16 | < 0.0001 |
| | C1NC | 5.21 | 0.36 | 14.63 | < 0.0001 |
| | C2C | 4.97 | 0.76 | 6.55 | < 0.0001 |
| | C2NC | 5.39 | 0.33 | 16.18 | < 0.0001 |
| | Visit | 0.01 | 0.11 | 0.10 | 0.9179 |
| | C1C*Visit | 0.44 | 0.34 | 1.29 | 0.249 |
| | C1NC*Visit | -0.81 | 0.18 | -4.42 | < 0.0001 |
| | C2C*Visit | -0.12 | 0.42 | -0.28 | 0.8629 |
| | C2NC*Visit | -0.92 | 0.17 | -5.53 | < 0.0001 |
| General | (Intercept) | 1.48 | 0.33 | 4.47 | < 0.0001 |
| | C1C | 9.08 | 0.92 | 9.87 | < 0.0001 |
| | C1NC | 9.11 | 0.51 | 17.91 | < 0.0001 |
| | C2C | 7.61 | 1.11 | 6.82 | < 0.0001 |
| | C2NC | 9.18 | 0.47 | 19.37 | < 0.0001 |
| | Visit | -0.16 | 0.16 | -0.97 | 0.4138 |
| | C1C*Visit | -0.35 | 0.52 | -0.66 | 0.5653 |
| | C1NC*Visit | -1.43 | 0.27 | -5.29 | < 0.0001 |
| | C2C*Visit | 0.16 | 0.66 | 0.24 | 0.8075 |
| | C2NC*Visit | -1.50 | 0.25 | -6.05 | < 0.0001 |

*Positive=SIPS: positive symptom sum score, Negative=SIPS: negative symptom sum score, Disorganization= SIPS disorganization symptom sum score, General=SIPS general symptom sum score, C1C=Cluster1 converter, C2C=Cluster2 converter, C1NC=Cluster1 non-converter, C2NC=Cluster2 non-converter, Std.err= Standard error.*

**Figure S3:** Longitudinal differences between clusters, converters, and healthy controls

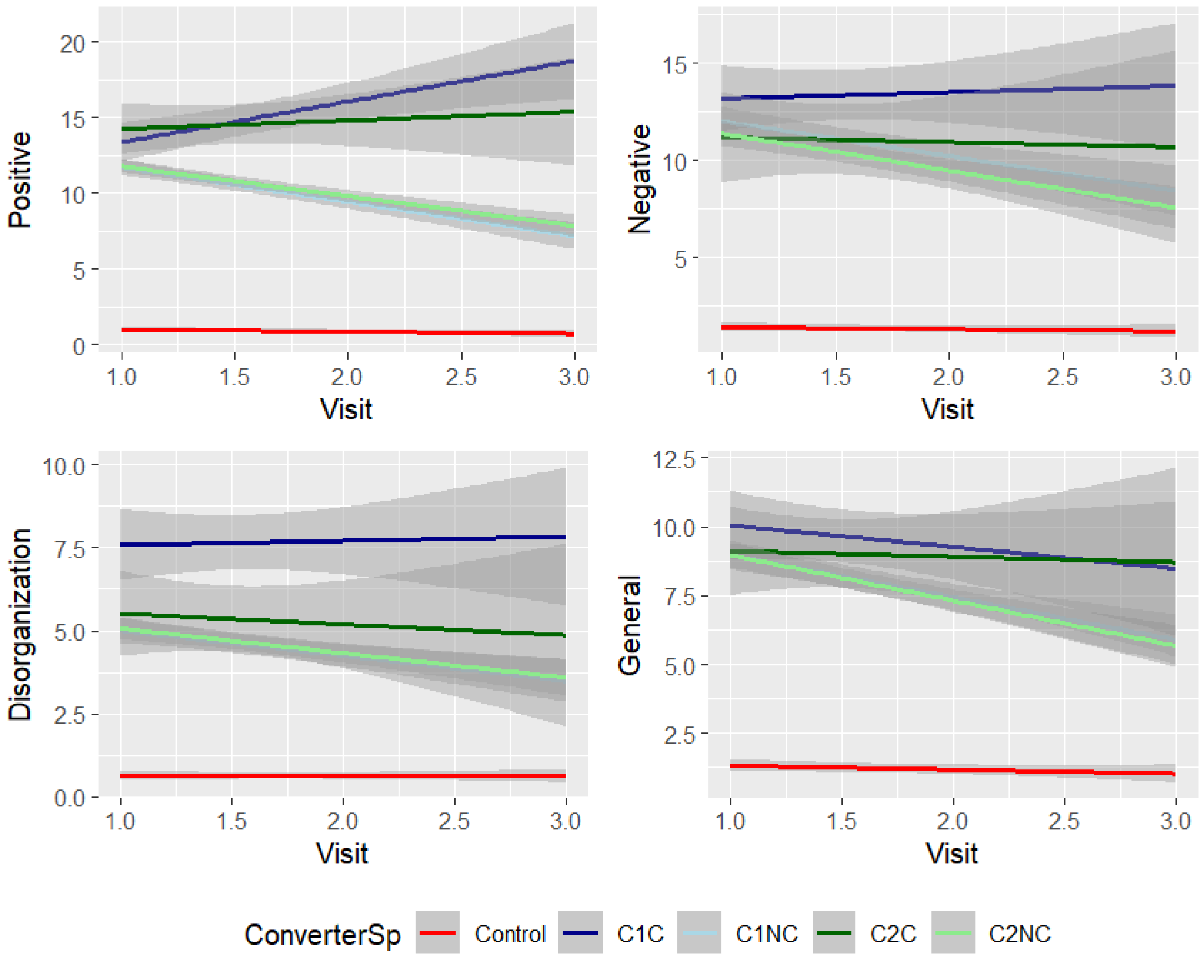

*Positive=SIPS: positive symptom sum score, Negative=SIPS: negative symptom sum score, Disorganization= SIPS disorganization symptom sum score, General=SIPS general symptom sum score, C1C=Cluster1 converter, C2C=Cluster2 converter, C1NC=Cluster1 non-converter, C2NC=Cluster2 non-converter*

**Table S6.** Relationship between cognition and SOPS as indicated by Generalized Estimating Equations

| Variable | Estimate | Std.err | Wald | p-value |
|---|---|---|---|---|
| Negative (CHR) | | | | |
| WRAT | -0.0326 | 0.0336 | 0.94 | 0.33 |
| Positive (Converters) | | | | |
| BACS | -0.09418 | 0.09898 | 0.91 | 1.000 |
| CPT Q3A | 0.16132 | 0.07309 | 4.87 | 0.189 |
| CPT QA | 0.07939 | 0.07645 | 1.08 | 0.350 |
| HVLT | 0.06553 | 0.09285 | 0.50 | 1.000 |
| LNS | -0.02883 | 0.08713 | 0.11 | 1.000 |
| WASI | 0.03377 | 0.09601 | 0.12 | 0.730 |
| WRAT | 0.02246 | 0.08851 | 0.06 | 1.000 |
| Positive (Non-converters) | | | | |
| BACS | -0.0385 | 0.0357 | 1.16 | 0.28 |
| CPT Q3A | -0.0725 | 0.0375 | 3.73 | 0.371 |
| CPT QA | -0.0598 | 0.0353 | 2.87 | 0.315 |
| HVLT | -0.0224 | 0.0383 | 0.34 | 0.98 |
| LNS | -0.0351 | 0.0385 | 0.83 | 0.84 |
| WASI | -0.0353 | 0.0345 | 1.05 | 0.362 |
| WRAT | -0.0122 | 0.0369 | 0.11 | 1 |
| Negative (Non-converters) | | | | |
| BACS | -0.1237 | 0.0368 | 11.27 | 0.00277** |
| CPT Q3A | -0.0920 | 0.0345 | 7.10 | 0.0134* |
| CPT QA | -0.1250 | 0.0327 | 14.58 | 0.00091*** |
| HVLT | -0.1172 | 0.0330 | 12.64 | 0.00089*** |
| LNS | -0.0880 | 0.0385 | 5.23 | 0.0308* |
| WASI | -0.0698 | 0.0381 | 3.36 | 0.0782 |
| WRAT | -0.0400 | 0.0371 | 1.16 | 0.28 |

| | | | | |
|---|---|---|---|---|
| Disorganization (Non-converters) | | | | |
| BACS | -0.0556 | 0.0400 | 1.93 | 0.3967 |
| CPT Q3A | -0.0654 | 0.0346 | 3.57 | 0.2065 |
| CPT QA | -0.1144 | 0.0297 | 14.9 | 0.00084*** |
| HVLT | -0.0081 | 0.0371 | 0.05 | 0.970 |
| LNS | -0.0297 | 0.0370 | 0.64 | 0.588 |
| WASI | 0.0115 | 0.0365 | 0.10 | 0.75 |
| WRAT | 0.0481 | 0.0368 | 1.71 | 0.3325 |
| General (Non-converters) | | | | |
| BACS | -0.0124 | 0.0370 | 0.11 | 0.74 |
| CPT Q3A | -0.0388 | 0.0340 | 1.30 | 0.25 |
| CPT QA | -0.0532 | 0.0311 | 2.92 | 0.088 |
| HVLT | -0.0413 | 0.0358 | 1.33 | 0.25 |
| LNS | -0.0203 | 0.0373 | 0.30 | 0.59 |
| WASI | 0.00236 | 0.03717 | 0.01 | 0.95 |
| WRAT | -0.0217 | 0.0367 | 0.35 | 0.55 |

*WRAT=Wide Range Achievement Test-Four Reading subtest, WASI Vocab=Wechsler Abbreviated Scale for Intelligence-2 Vocabulary, CPT= auditory working memory continuous performance test; QA=vigilance;Q3A-MEM= working memory load/no interference, BACS=Brief Assessment of Cognition in Schizophrenia-symbol coding, HVLT-R= Hopkins Verbal Learning Test-Revised, LNS= Letter-number-span, Positive=SIPS: positive symptom sum score, Negative=SIPS: negative symptom sum score, Disorganization= SIPS disorganization symptom sum score, General=SIPS general symptom sum score, Std.err= Standard error.*

**Figure S4:** Survival Curves for the CHR conversion by cluster

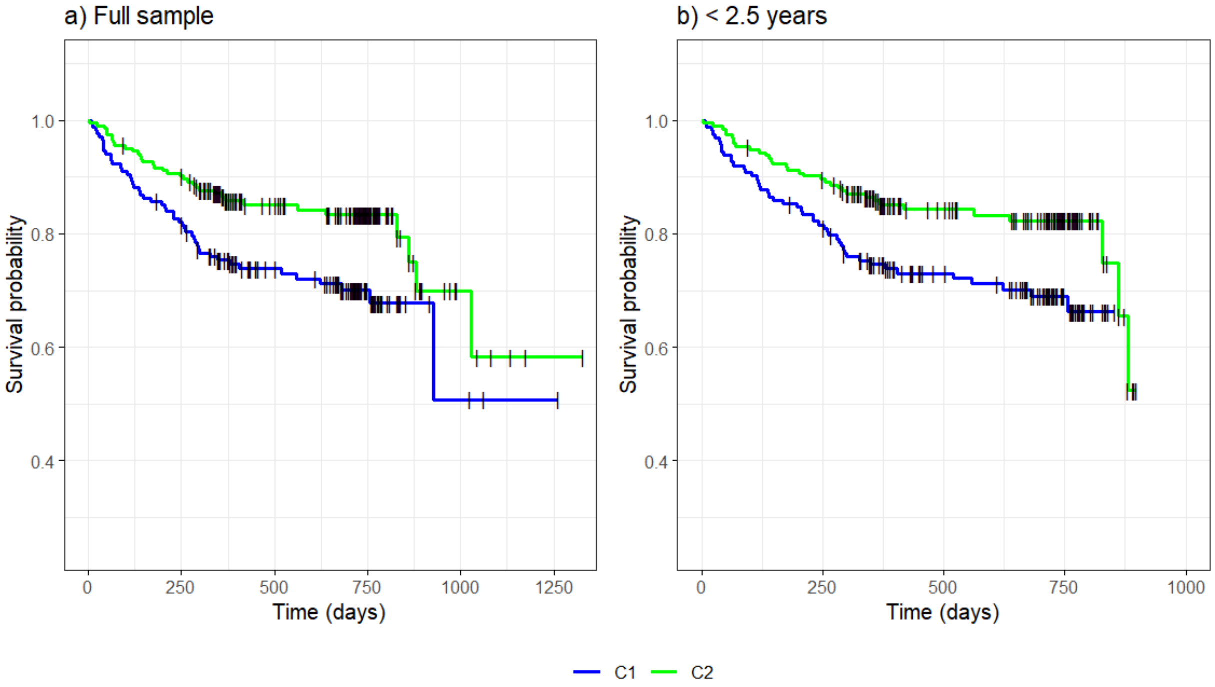

*Vertical black dashes indicate censored data. C=Cluster*

**Table S7.** Relationship between cognition and functioning as indicated by Generalized Estimating Equations

| Cognition (Predictor) | Functioning (Response) | Estimate | Std.err | Wald | p-value |
|---|---|---|---|---|---|
| BACS | RS | 0.2440 | 0.0252 | 94.10 | <2e-16 |
| | SS | 0.1908 | 0.0261 | 53.39 | 8.1e-13 |
| | GAF | 0.2233 | 0.0239 | 87.03 | <2e-16 |
| CPT Q3A | RS | 0.0700 | 0.0231 | 9.22 | 0.0072 |
| | SS | 0.0975 | 0.0216 | 20.30 | 1.9e-05 |
| | GAF | 0.0768 | 0.0190 | 16.41 | 15e-04 |
| CPT QA | RS | 0.0647 | 0.0230 | 7.88 | 0.015 |
| | SS | 0.0956 | 0.0212 | 20.40 | 1.8e-05 |
| | GAF | 0.0929 | 0.0231 | 16.14 | 17e-04 |
| HVLT | RS | 0.2112 | 0.0235 | 81.11 | 2.34e-15 |
| | SS | 0.1207 | 0.0217 | 31.03 | 7.5e-8 |
| | GAF | 0.1392 | 0.0213 | 42.58 | 2.04e-10 |
| LNS | RS | 0.2208 | 0.0258 | 73.26 | <2e-16 |
| | SS | 0.1783 | 0.0255 | 48.76 | 8.7e-12 |
| | GAF | 0.1819 | 0.0246 | 54.81 | 3.9e-13 |
| WASI | RS | 0.2549 | 0.0246 | 106.94 | <2e-16 |
| | SS | 0.1496 | 0.0271 | 30.6 | 9.6e-8 |
| | GAF | 0.2024 | 0.0246 | 67.70 | 6.6e-16 |
| WRAT | RS | 0.1334 | 0.0259 | 26.47 | 8.1e-7 |
| | SS | 0.0944 | 0.0236 | 15.99 | 19e-04 |
| | GAF | 0.0979 | 0.0214 | 20.98 | 1.3e-06 |

*WRAT=Wide Range Achievement Test-Four Reading subtest, WASI Vocab=Wechsler Abbreviated Scale for Intelligence-2 Vocabulary, CPT= auditory working memory continuous performance test; QA=vigilance;Q3A-MEM= working memory load/no interference, BACS=Brief Assessment of Cognition in Schizophrenia-symbol coding, HVLT-R= Hopkins Verbal Learning*

*Test-Revised, LNS= Letter-number-span, GAF= Global Assessment of Functioning, GF:SS= Global Functioning social scale, GF:RS= Global Functioning social scale role scales, Std.err= Standard error.*

**Table S8 .** Relationship between cognition and functioning as indicated by Generalized Estimating Equations

| Cognition (Response) | Functioning (Predictor) | Estimate | Std.err | Wald | p-value |
|---|---|---|---|---|---|
| BACS | RS | 0.111 | 0.024 | 20.01 | 7.7e-06 |
| | SS | 0.115 | 0.026 | 19.4 | 1.1e-05 |
| | GAF | 0.198 | 0.024 | 64.7 | 8.9e-16 |
| CPT Q3A | RS | 0.09 | 0.024 | 15.8 | 7e-05 |
| | SS | 0.127 | 0.025 | 26.07 | 3.3e-05 |
| | GAF | 0.2 | 0.024 | 69.77 | <2e-16 |
| CPT QA | RS | 0.126 | 0.023 | 28.35 | 1e-07 |
| | SS | 0.158 | 0.03 | 27.63 | 1.5e-07 |
| | GAF | 0.188 | 0.027 | 46.2 | 1e-11 |
| HVLT | RS | 0.226 | 0.026 | 75.63 | <2e-16 |
| | SS | 0.169 | 0.025 | 44.95 | 2e-11 |
| | GAF | 0.232 | 0.026 | 75.2 | <2e-16 |
| LNS | RS | 0.164 | 0.023 | 48.61 | 3.1e-12 |
| | SS | 0.165 | 0.025 | 42.32 | 7.7e-11 |
| | GAF | 0.209 | -.026 | 61.53 | 4.3e-15 |
| WASI | RS | 0.184 | 0.022 | 65.86 | 4.4e-16 |
| | SS | 0.129 | 0.024 | 27.83 | 1.3e-07 |
| | GAF | 0.207 | 0.024 | 71.19 | <2e-16 |
| WRAT | RS | 0.098 | 0.023 | 17.91 | 2.3e-05 |
| | SS | 0.095 | 0.024 | 15.8 | 7e-05 |
| | GAF | 0.127 | 0.023 | 29.43 | 5.8e-08 |

*WRAT=Wide Range Achievement Test-Four Reading subtest, WASI Vocab=Wechsler Abbreviated Scale for Intelligence-2 Vocabulary, CPT= auditory working memory continuous performance test; QA=vigilance;Q3A-MEM= working memory load/no interference, BACS=Brief Assessment of Cognition in Schizophrenia-symbol coding, HVLT-R= Hopkins Verbal Learning Test-Revised, LNS= Letter-number-span, GAF= Global Assessment of Functioning, GF:SS= Global Functioning social scale, GF:RS= Global Functioning social scale role scales, Std.err= Standard error.*
